# Estimated global and regional incidence and prevalence of herpes simplex virus infections and genital ulcer disease in 2020: Mathematical modeling analyses

**DOI:** 10.1101/2024.06.03.24308350

**Authors:** Manale Harfouche, Sawsan AlMukdad, Asalah Alareeki, Aisha M. M. Osman, Sami L. Gottlieb, Jane Rowley, Laith J. Abu-Raddad, Katharine J. Looker

## Abstract

**Background:** Genital herpes simplex virus (HSV) type 1 and 2 infections are lifelong and can cause symptomatic genital ulcer disease (GUD). HSV-2 almost always causes sexually transmitted genital infection, while HSV-1 mainly causes oral infection but can be sexually transmitted to cause genital infection. This study estimated genital infection with both HSV types and associated GUD globally in 2020, breaking down the data by World Health Organization (WHO) region and sex for females and males.

**Methods:** A calibrated mathematical model was employed to generate estimates for the incidence and prevalence of HSV-2 infection, genital HSV-1 infection, and GUD caused by both HSV types. Estimates for non-genital infections caused by HSV-1 were also generated. Model input was derived from a comprehensive systematic review and meta-analyses of HSV prevalence data for all WHO regions.

**Results:** Globally in 2020 there were 25.6 million (95% uncertainty interval (UI): 23.1-29.4 million) people aged 15 to 49 years with new HSV-2 infections and 519.5 million (95% UI: 464.3-611.3 million), or 13.3% (95% UI: 11.9-15.6%), with existing (prevalent) HSV-2 infections. In addition, there were 16.8 million (95% UI: 10.6-22.4 million) people aged 15-49 years with new genital HSV-1 infections and 376.2 million (95% UI: 235.6-483.5 million), or 10.2% (95% UI: 6.4-13.1%), with prevalent genital HSV-1 infections. The estimated number of people aged 15 to 49 years with at least one episode of HSV-attributable genital ulcer disease in 2020 was 187.9 million (95% UI: 116.0-291.8 million) for HSV-2 and 16.7 million (95% UI: 9.3-25.2 million) for HSV-1, totaling 204.6 million (95% UI: 132.3-306.5).

**Conclusion:** Genital HSV infections have a high incidence and prevalence worldwide, contributing to a significant GUD disease burden. New prevention and treatment measures, such as prophylactic and therapeutic HSV vaccines, are critically needed to control HSV infections and reduce the associated disease burden.

## Introduction

Herpes simplex virus (HSV) type 1 and type 2 cause lifelong infections and are widespread worldwide, resulting in significant disease and economic burden [1–3]. Both infections are characterized by frequent infectious subclinical shedding, along with symptomatic reactivations in a proportion of those infected [4–8]. HSV-2 infection is almost always acquired through sexual transmission and is the leading cause of recurrent clinical genital ulcer disease (GUD) in most countries [9–12]. In addition to the painful genital sores, genital herpes is associated with a range of social and psychological adverse outcomes, including effects on sexual relations, quality of life, and mental health issues such as depression, anxiety, and low self-esteem [13–16]. HSV-2 infection is believed to increase the risk of HIV acquisition and transmission by threefold [17, 18], potentially resulting in an epidemiological synergy between these two infections [18–20].

HSV-1 infection is typically acquired orally in childhood and commonly manifests as cold sores [21, 22], but it can also cause more serious neurological, corneal, and mucocutaneous complications [23–25]. Adults can acquire genital HSV-1 infection if they were not infected orally during childhood [26]. Particularly in high-income countries, HSV-1 infection has been increasingly acquired sexually [9, 26–28], and in several countries, it is now the leading cause of first-episode genital herpes [9, 27, 28]. Recurrences of genital HSV-1 are much less frequent compared to HSV-2 recurrences [4].

HSV-1 and HSV-2 infections can both be transmitted, albeit rarely, from genitally-infected mothers to their neonates during birth and through oral contact from caregivers postnatally [29], leading to neonatal herpes, a disabling disease in newborns with a high fatality rate [14, 30]. In response to the clinical disease burden of these two infections and their impact on sexual and reproductive health and HIV transmission, the World Health Organization (WHO) has advocated for efforts to reduce HSV disease burden by advancing the development of new prevention and treatment measures, such as vaccines [31].

In this article, we present global and regional modeling estimates of the incidence and prevalence of genital HSV infections as well as HSV-related genital ulcer disease (GUD) for the year 2020. In addition to updating earlier estimates [1, 32, 33], the present estimates benefit from methodological enhancements. The main enhancement is that HSV-1 and HSV-2 prevalence data, which provided the modeling input, are based on a series of standardized and comprehensive HSV systematic reviews, meta-analyses, and meta-regressions covering all global regions, published over the last few years [9-12, 27, 28, 34-40].

## Methods

### Model input

The model input was provided by the databases of the published systematic reviews [9-12, 27, 28, 34-40]. Since the reviews were published over several years [9-12, 27, 28, 34-40], and considering the lag of a few years between sample collection and data publication [9-12, 27, 28, 34-40], an update was done up to March 30, 2022, to ensure completeness and inclusion of the most recent data for each WHO region. However, only data based on samples collected up to 2020 were used for the modeling input. Since the regional groupings of countries in the published reviews were not WHO region groupings, country-level data were re-grouped into the six WHO regions. While the details of the standardized methodology of the reviews have been published previously [9-12, 27, 28, 34-40], a summary is provided in Box S1 of the Supplementary Material for inclusiveness.

The validity of each HSV diagnostic assay for each data point was investigated, considering known limitations of HSV serology and potential cross-reactivity between HSV-1 and HSV-2 antibodies [41, 42]. This assessment was conducted in consultation with Professor Rhoda Ashley-Morrow of the University of Washington, an expert in HSV diagnostic methods. Only studies with valid and reliable assays were included in the systematic reviews.

Data were synthesized based on methods used to generate the 2016 WHO HSV estimates [1], but with adjustments to improve on study methodology, to incorporate recent findings on HSV-1 and HSV-2 epidemiology, and to account for limitations in available data for specific regions (Box S1). These adjustments are summarized in Box S2. Inclusion criteria for HSV-1 was an antibody prevalence (seroprevalence) measure in a general population aged 0-49 years with a midpoint of 2004 up to 2020 for the year of data collection (and 1995 up to 2020 for the African and South-East Asia regions due to poor data availability). Inclusion criteria for HSV-2 was an antibody prevalence measure among a general population aged 15-49 years with a midpoint of 2004 up to 2020 for the year of data collection.

Meta-analyses were conducted using the DerSimonian-Laird random-effects models [43] with the Freeman-Tukey double arcsine transformation [44]. Pooled mean HSV-1 and HSV-2 prevalences were calculated by sex (when possible) and by age. In this simplified representation of populations, we stratified populations into two groups, “females” and “males”; however, we recognize that a range of gender identities are possible. The meta package [45] was used to perform the meta-analyses in R, version 4.0.4 [46] (Box S1). The pooled prevalence estimates for each region were subsequently used to calibrate the mathematical model [1, 32, 33] that generated the HSV estimations.

### Model calibrations

HSV-1 and HSV-2 incidence estimates were calibrated to the pooled mean prevalences using maximum likelihood, as per the previously published WHO HSV estimation model [1]. Any prevalence values of 0% and 100% were recoded as 0.1% and 99.9%, respectively, before taking logs for the likelihood equations. For the calibration, a constant force of infection, *λ*, was assumed; that is, assuming a constant incidence rate model. An additional term, *k*, representing the maximum proportion of individuals that can be infected, was also included and simultaneously calibrated along with *λ*. It was assumed that individuals can be infected with HSV-1 from age 0 years, and with HSV-2 from age 15 years. Calibration was done using the Solver function in Microsoft Excel. Further details and the model equations can be found in Box S3.

For HSV-1, single data points were included in the calibrations for the WHO South-East Asia region, due to particularly poor data availability. Sex-specific estimates were produced for the Americas and European regions as conducted previously [1], but for children (<15 years), all of female, male, and mixed data for a given age band were pooled and used for calibration for each of females and males, to avoid skewing the model calibrations by sex. For the Western Pacific, African, Eastern Mediterranean, and South-East Asia regions all of female, male, and mixed data were pooled for a given age band. Previous meta-regressions on all prevalence data for these regions showed no difference in prevalence by sex whether for Africa [38], Eastern Mediterranean [36], and Asia (combining South-East Asia and Western Pacific regions) [39]. For HSV-2, single data points were included in the calibration for the Eastern Mediterranean region, again due to poor data availability.

Despite the steps taken to maximize the use of available data, data were still sparse for some calibrations. The model fit for the Eastern Mediterranean region was very skewed due to low availability of HSV-1 related data among children. A better fit for this region was produced by using the data for children from the Western Pacific region, due to close similarity in pooled prevalence for HSV-1 among adults in these two regions.

### Prevalence and incidence estimates

Smoothed prevalence estimates by sex and 5-year age group and calibrated incidence obtained from the model fitting were then applied to demographic data by sex and 5-year age group for 2020 obtained from the United Nations Population Division [47].

Genital infections with HSV-1 were assumed to occur only in individuals over the age of 15. All individuals under 15 years of age were assumed to have acquired the infection orally. Among individuals over 15 years of age, we used the same pooled values as in the 2016 estimates: 72.4% of new infections over 15 years of age were assumed to be genital, and 36.4% oral infections (percentages add up to more than 100% due to dual oral and genital infections) [1].

Estimates for genital HSV-1 and HSV-2 infections were generated for individuals aged 15-49 years. The percentage of individuals with genital infection due to either HSV-1 or HSV-2 was calculated by summing the separate estimates for genital HSV-1 and HSV-2, and then applying a correction factor for the estimated percentage with dual HSV-1 and HSV-2 infection (Box S3).

Estimates for oral HSV-1 infection were generated for individuals aged 0-49 years.

As in the 2016 estimation round [1], a secondary exploratory analysis was conducted where the number of individuals with HSV-1 and HSV-2 infections in each WHO region for those aged 50-99 years was estimated by applying the prevalence of each infection in individuals aged 45-49 years to the population aged 50-99 years. Calculation of all estimates was done in Excel.

### Uncertainty analysis

Similar to the 2016 estimates [1], 95% uncertainty intervals (UI) were computed. In brief, the force of infection, *λ*, was sampled 1000 times in Excel (keeping *k* constant), with standard error derived from values for *λ* calibrated to the lower and upper 95% confidence intervals (CI) for the pooled mean prevalences, while assuming a normal distribution. The percentages of individuals from age 15 years that are infected orally versus genitally were simultaneously sampled 1000 times. The 95% UIs were then derived from these 1000 runs.

### GUD estimates

GUD due to HSV-2 or genital HSV-1 infection was estimated using two measures following our published method: [2] (1) the percentage and the number of people aged 15-49 years with any HSV GUD in 2020; and (2) the total number of person-days with HSV GUD among individuals aged 15-49 years old in 2020. GUD estimates were calculated by applying natural history estimates for HSV-2 and genital HSV-1 infections, i.e., the probability and duration of a first episode, the probability of a recurrence, and the length and frequency of recurrences, to the HSV-2 and HSV-1 infection estimates for 2020. These natural history estimates were derived by pooling study-level estimates obtained from a comprehensive literature review [2]. It was assumed that the total HSV-related GUD burden was equal to the sum of the burden for each of HSV-2 and genital HSV-1. Calculation of all estimates was done in Excel. Further details of how to obtain these estimates have been previously published [2].

The 95% UI for the percentage and number of individuals aged 15-49 years with any GUD and the total number of person-days with GUD among 15-49 years old individuals were derived as for 2016 [2]. In brief, all natural history parameters, and the calibrated force of infection (as above), were sampled 1000 times in Excel. The 95% UIs were then derived from these 1000 runs.

### Compliance with guidelines

This study complies with the Guidelines for Accurate and Transparent Health Estimates Reporting (GATHER) recommendation [48]. The complete GATHER checklist can be found in Table S1 of the Supplementary Material.

## Results

Given that HSV-2 is the primary contributor to genital infection and recurrent GUD, the results are presented first for HSV-2.

### Model input and fitting

In the published reviews [9-12, 27, 28, 34-40] and their update to complete the database of HSV data used for the model input, titles and abstracts of 76,972 citations were screened for relevant HSV-2 and HSV-1 data for all WHO regions. Of these, 1,228 articles reported an HSV-2 and/or HSV-1 epidemiological outcome measures (Table S2). A total of 134 articles included data that met the specific inclusion criteria for the 2020 HSV-2 estimates (list of articles in Box S4) and 82 articles included data that met the specific inclusion criteria for the 2020 HSV-1 estimates (list of articles in Box S5).

In comparison to the 2016 estimates [1], the number of available data points improved for most of the regions (Tables S3, S4, and S5). Globally, the number of articles in 2020 surpassed that of 2016. Specifically, the number of articles on HSV-2 increased from 88 to 134, and for HSV-1, it rose from 44 to 82 (Table S5). However, this increased availability did not necessarily translate into a larger number of represented countries in some of the regions.

Figure S1 shows the model fits for HSV-2 prevalence versus age for each WHO region. Overall, the model fitted well available data. These model fits were subsequently used to generate HSV-2 incidence and prevalence estimates.

Figure S2 shows the model fits for HSV-1 prevalence versus age for each WHO region. Overall, the model fitted well available data. These model fits were subsequently used to generate HSV-1 incidence and prevalence estimates.

### HSV-2 infection

The number of incident HSV-2 infections globally in 2020 among individuals aged 15-49 years was estimated at 25.6 million (95% UI: 23.1-29.4 million) (Table 1). Of these, 15.6 million (95% UI: 13.8-18.0 million) infections were among females and 10.0 million (95% UI: 8.4-12.9 million) among males. The African region had the highest HSV-2 incidence for both females and males at nearly 10 million infections, accounting for 38.3% of all infections. HSV-2 incidence rate decreased with age for all regions and was notably high among young adults in the African and Americas regions.

**Table 1.**
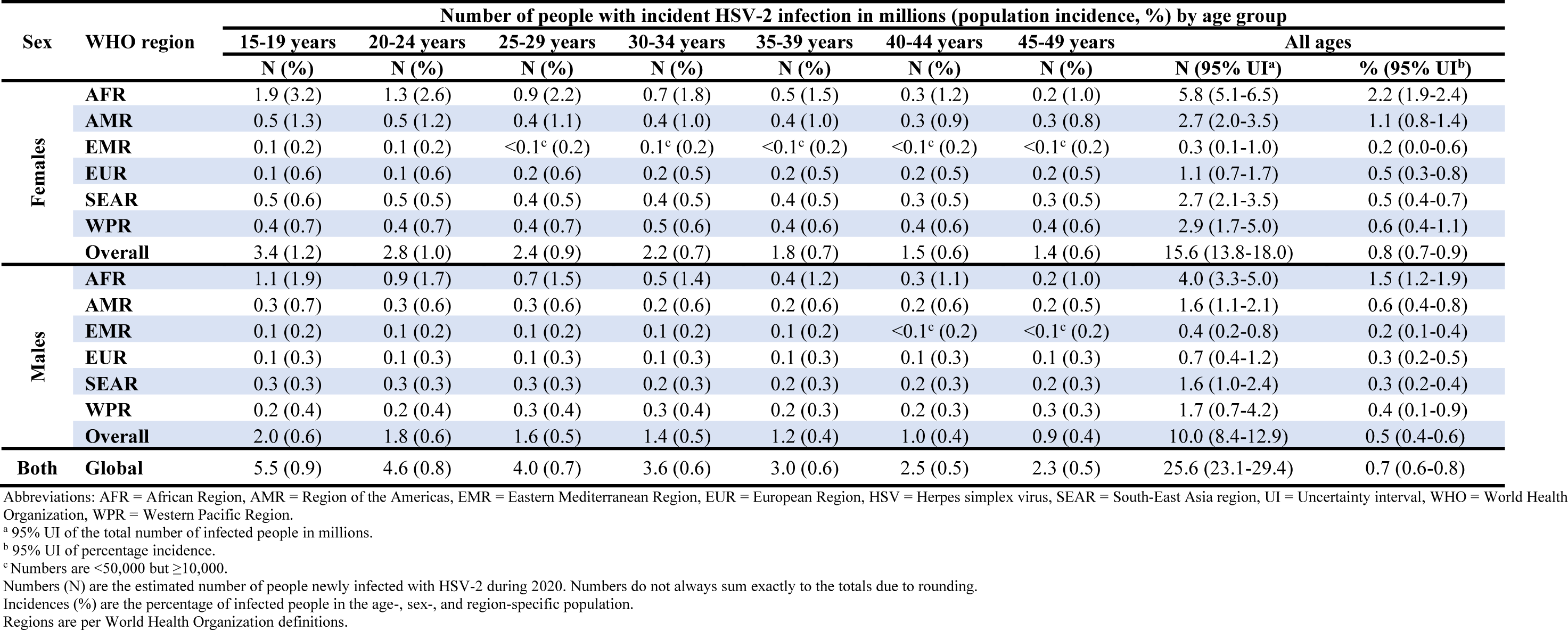
Global and regional estimates of the number and percentage of the population aged 15-49 years with incident HSV-2 infection in 2020, by age and sex.

| Sex | WHO region | Number of people with incident HSV-2 infection in millions (population incidence, %) by age group |  |  |  |  |  |  |  |  |
| --- | --- | --- | --- | --- | --- | --- | --- | --- | --- | --- |
|  |  | 15-19 years | 20-24 years | 25-29 years | 30-34 years | 35-39 years | 40-44 years | 45-49 years | All ages |  |
|  |  | N (%) | N (%) | N (%) | N (%) | N (%) | N (%) | N (%) | N (95% UI<sup>a</sup>) | % (95% UI<sup>b</sup>) |
| Females | AFR | 1.9 (3.2) | 1.3 (2.6) | 0.9 (2.2) | 0.7 (1.8) | 0.5 (1.5) | 0.3 (1.2) | 0.2 (1.0) | 5.8 (5.1-6.5) | 2.2 (1.9-2.4) |
|  | AMR | 0.5 (1.3) | 0.5 (1.2) | 0.4 (1.1) | 0.4 (1.0) | 0.4 (1.0) | 0.3 (0.9) | 0.3 (0.8) | 2.7 (2.0-3.5) | 1.1 (0.8-1.4) |
|  | EMR | 0.1 (0.2) | 0.1 (0.2) | <0.1<sup>c</sup> (0.2) | 0.1<sup>c</sup> (0.2) | <0.1<sup>c</sup> (0.2) | <0.1<sup>c</sup> (0.2) | <0.1<sup>c</sup> (0.2) | 0.3 (0.1-1.0) | 0.2 (0.0-0.6) |
|  | EUR | 0.1 (0.6) | 0.1 (0.6) | 0.2 (0.6) | 0.2 (0.5) | 0.2 (0.5) | 0.2 (0.5) | 0.2 (0.5) | 1.1 (0.7-1.7) | 0.5 (0.3-0.8) |
|  | SEAR | 0.5 (0.6) | 0.5 (0.5) | 0.4 (0.5) | 0.4 (0.5) | 0.4 (0.5) | 0.3 (0.5) | 0.3 (0.5) | 2.7 (2.1-3.5) | 0.5 (0.4-0.7) |
|  | WPR | 0.4 (0.7) | 0.4 (0.7) | 0.4 (0.7) | 0.5 (0.6) | 0.4 (0.6) | 0.4 (0.6) | 0.4 (0.6) | 2.9 (1.7-5.0) | 0.6 (0.4-1.1) |
|  | Overall | 3.4 (1.2) | 2.8 (1.0) | 2.4 (0.9) | 2.2 (0.7) | 1.8 (0.7) | 1.5 (0.6) | 1.4 (0.6) | 15.6 (13.8-18.0) | 0.8 (0.7-0.9) |
| Males | AFR | 1.1 (1.9) | 0.9 (1.7) | 0.7 (1.5) | 0.5 (1.4) | 0.4 (1.2) | 0.3 (1.1) | 0.2 (1.0) | 4.0 (3.3-5.0) | 1.5 (1.2-1.9) |
|  | AMR | 0.3 (0.7) | 0.3 (0.6) | 0.3 (0.6) | 0.2 (0.6) | 0.2 (0.6) | 0.2 (0.6) | 0.2 (0.5) | 1.6 (1.1-2.1) | 0.6 (0.4-0.8) |
|  | EMR | 0.1 (0.2) | 0.1 (0.2) | 0.1 (0.2) | 0.1 (0.2) | 0.1 (0.2) | <0.1<sup>c</sup> (0.2) | <0.1<sup>c</sup> (0.2) | 0.4 (0.2-0.8) | 0.2 (0.1-0.4) |
|  | EUR | 0.1 (0.3) | 0.1 (0.3) | 0.1 (0.3) | 0.1 (0.3) | 0.1 (0.3) | 0.1 (0.3) | 0.1 (0.3) | 0.7 (0.4-1.2) | 0.3 (0.2-0.5) |
|  | SEAR | 0.3 (0.3) | 0.3 (0.3) | 0.3 (0.3) | 0.2 (0.3) | 0.2 (0.3) | 0.2 (0.3) | 0.2 (0.3) | 1.6 (1.0-2.4) | 0.3 (0.2-0.4) |
|  | WPR | 0.2 (0.4) | 0.2 (0.4) | 0.3 (0.4) | 0.3 (0.4) | 0.2 (0.3) | 0.2 (0.3) | 0.3 (0.3) | 1.7 (0.7-4.2) | 0.4 (0.1-0.9) |
|  | Overall | 2.0 (0.6) | 1.8 (0.6) | 1.6 (0.5) | 1.4 (0.5) | 1.2 (0.4) | 1.0 (0.4) | 0.9 (0.4) | 10.0 (8.4-12.9) | 0.5 (0.4-0.6) |
| Both | Global | 5.5 (0.9) | 4.6 (0.8) | 4.0 (0.7) | 3.6 (0.6) | 3.0 (0.6) | 2.5 (0.5) | 2.3 (0.5) | 25.6 (23.1-29.4) | 0.7 (0.6-0.8) |

Abbreviations: AFR = African Region, AMR = Region of the Americas, EMR = Eastern Mediterranean Region, EUR = European Region, HSV = Herpes simplex virus, SEAR = South-East Asia region, UI = Uncertainty interval, WHO = World Health Organization, WPR = Western Pacific Region.
<sup>a</sup> 95% UI of the total number of infected people in millions.
<sup>b</sup> 95% UI of percentage incidence.
<sup>c</sup> Numbers are <50,000 but ≥10,000.
Numbers (N) are the estimated number of people newly infected with HSV-2 during 2020. Numbers do not always sum exactly to the totals due to rounding.
Incidences (%) are the percentage of infected people in the age-, sex-, and region-specific population.
Regions are per World Health Organization definitions.

The number of people with prevalent HSV-2 infections globally in 2020 among individuals aged 15-49 years was estimated at 519.5 million (95% UI: 464.3-611.3 million), an HSV-2 prevalence of 13.3% (95% UI: 11.9-15.6%) (Table 2). HSV-2 prevalence was higher in females (17.0%; 95% UI: 14.9-20.1%) than in males (9.7%; 95% UI: 8.0-13.0%). The African region exhibited both the highest prevalence and the largest number of infected persons. In the secondary analysis, among individuals aged 50-99 years, an additional 389.1 million people were estimated to be infected worldwide (Table S6).

**Table 2.** Global and regional estimates of the number and percentage of the population aged 15-49 years with prevalent HSV-2 infection in 2020, by age and sex.

| Sex | WHO region | Number of people with prevalent HSV-2 infection in millions (population prevalence, %) by age group |  |  |  |  |  |  |  |  |
| --- | --- | --- | --- | --- | --- | --- | --- | --- | --- | --- |
|  |  | 15-19 years | 20-24 years | 25-29 years | 30-34 years | 35-39 years | 40-44 years | 45-49 years | All ages |  |
|  |  | N (%) | N (%) | N (%) | N (%) | N (%) | N (%) | N (%) | N (95% UI<sup>a</sup>) | % (95% UI<sup>b</sup>) |
| Females | AFR | 10.3 (19.3) | 16.2 (33.6) | 19.2 (45.3) | 20.4 (55.0) | 20.0 (63.0) | 18.0 (69.5) | 15.7 (74.9) | 119.8 (99.8-141.8) | 44.4 (37.0-52.5) |
|  | AMR | 2.5 (7.2) | 4.9 (13.4) | 7.3 (19.1) | 9.1 (24.4) | 10.6 (29.4) | 11.4 (34.1) | 12.0 (38.4) | 57.8 (42.2-77.4) | 22.6 (16.5-30.2) |
|  | EMR | 0.3 (0.9) | 0.5 (1.8) | 0.7 (2.6) | 0.9 (3.4) | 1.0 (4.2) | 1.0 (5.0) | 1.0 (5.8) | 5.5 (1.4-19.1) | 3.0 (0.8-10.5) |
|  | EUR | 0.8 (3.3) | 1.5 (6.1) | 2.5 (8.9) | 3.8 (11.6) | 4.7 (14.3) | 5.4 (16.8) | 6.1 (19.3) | 24.7 (15.2-39.3) | 11.7 (7.2-18.6) |
|  | SEAR | 2.4 (3.1) | 4.7 (5.8) | 6.6 (8.4) | 8.4 (11.0) | 9.8 (13.5) | 10.4 (15.9) | 10.8 (18.3) | 53.1 (40.2-69.1) | 10.0 (7.6-13.0) |
|  | WPR | 2.0 (4.0) | 4.1 (7.5) | 6.7 (10.8) | 10.9 (14.0) | 11.1 (17.2) | 12.5 (20.2) | 17.1 (23.1) | 64.5 (35.8-116.4) | 14.0 (7.8-25.3) |
|  | Overall | 18.2 (6.2) | 32.0 (11.1) | 43.2 (15.0) | 53.5 (18.2) | 57.1 (21.4) | 58.7 (24.2) | 62.7 (26.5) | 325.5 (284.9-383.0) | 17.0 (14.9-20.1) |
| Males | AFR | 5.8 (10.7) | 9.5 (19.5) | 11.6 (27.4) | 12.7 (34.5) | 12.7 (41.0) | 11.8 (46.8) | 10.6 (52.0) | 74.8 (59.0-97.0) | 27.7 (21.9-36.0) |
|  | AMR | 1.3 (3.7) | 2.7 (7.0) | 4.0 (10.2) | 4.9 (13.2) | 5.7 (16.2) | 6.2 (19.0) | 6.5 (21.8) | 31.3 (22.1-43.3) | 12.2 (8.6-16.9) |
|  | EMR | 0.3 (1.1) | 0.6 (2.0) | 0.9 (3.0) | 1.2 (4.0) | 1.3 (4.9) | 1.3 (5.8) | 1.2 (6.8) | 7.0 (3.0-16.0) | 3.5 (1.5-8.1) |
|  | EUR | 0.4 (1.8) | 0.9 (3.4) | 1.5 (5.0) | 2.2 (6.5) | 2.6 (8.0) | 3.0 (9.5) | 3.4 (11.0) | 14.0 (7.6-24.9) | 6.5 (3.5-11.6) |
|  | SEAR | 1.4 (1.6) | 2.7 (3.1) | 3.9 (4.6) | 4.8 (6.0) | 5.6 (7.4) | 5.9 (8.8) | 6.2 (10.2) | 30.6 (19.7-46.3) | 5.4 (3.5-8.2) |
|  | WPR | 1.1 (2.1) | 2.4 (3.9) | 3.9 (5.7) | 6.1 (7.5) | 6.2 (9.2) | 7.0 (10.9) | 9.6 (12.6) | 36.5 (13.8-93.3) | 7.4 (2.8-18.9) |
|  | Overall | 10.5 (3.3) | 18.8 (6.1) | 25.8 (8.5) | 32.0 (10.4) | 34.3 (12.5) | 35.3 (14.2) | 37.5 (15.6) | 194.1 (160.0-260.5) | 9.7 (8.0-13.0) |
| Both | Global | 28.7 (4.7) | 50.8 (8.5) | 69.0 (11.7) | 85.5 (14.2) | 91.4 (16.9) | 94.0 (19.2) | 100.2 (21.0) | 519.5 (464.3-611.3) | 13.3 (11.9-15.6) |

Abbreviations: AFR = African Region, AMR = Region of the Americas, EMR = Eastern Mediterranean Region, EUR = European Region, HSV = Herpes simplex virus, SEAR = South-East Asia region, UI = Uncertainty interval, WHO = World Health Organization, WPR = Western Pacific Region.
<sup>a</sup> 95% UI of the total number of infected people in millions.
<sup>b</sup> 95% UI of percentage prevalence.
Numbers (N) are the year 2020 estimated number of people living with HSV-2 infection. Numbers do not always sum exactly to the totals due to rounding.
Prevalences (%) are the percentage of infected people in the age-, sex-, and region-specific population.
Regions are per World Health Organization definitions.

### Genital HSV-1 infection

The number of incident genital HSV-1 infections globally in 2020 among individuals aged 15-49 years was estimated at 16.8 million (95% UI: 10.6-22.4 million) (Table 3). Of these, 8.4 million (95% UI: 5.1-11.5 million) infections were among females and 8.4 million (95% UI: 5.0-11.8 million) among males. The Western Pacific region had the highest incidence of genital HSV-1, with 4.8 million infections combining females and males. Genital HSV-1 incidence rate decreased with age for all regions and was notably high among young adults in the Americas and European regions.

**Table 3.** Global and regional estimates of the number and percentage of the population aged 15-49 years with incident genital HSV-1 infection in 2020, by age and sex.

| Sex | WHO region | Number of people with incident genital HSV-1 infection in millions (population incidence, %) by age group |  |  |  |  |  |  |  |  |
| --- | --- | --- | --- | --- | --- | --- | --- | --- | --- | --- |
|  |  | 15-19 years | 20-24 years | 25-29 years | 30-34 years | 35-39 years | 40-44 years | 45-49 years | All ages |  |
|  |  | N (%) | N (%) | N (%) | N (%) | N (%) | N (%) | N (%) | N (95% UI<sup>a</sup>) | % (95% UI<sup>b</sup>) |
| Females | AFR | <0.1<sup>c</sup> (0.1) | <0.1<sup>d</sup> (0.0) | <0.1<sup>d</sup> (0.0) | 0.0 (0.0) | 0.0 (0.0) | 0.0 (0.0) | 0.0 (0.0) | 0.0 (0.0-0.5) | 0.0 (0.0-0.3) |
|  | AMR | 0.5 (1.4) | 0.4 (1.1) | 0.4 (0.9) | 0.3 (0.8) | 0.2 (0.6) | 0.2 (0.5) | 0.1 (0.4) | 2.1 (1.4-2.6) | 0.9 (0.6-1.0) |
|  | EMR | 0.2 (0.7) | 0.1 (0.3) | <0.1<sup>c</sup> (0.2) | <0.1<sup>c</sup> (0.1) | <0.1<sup>c</sup> (0.0) | <0.1<sup>d</sup> (0.0) | <0.1<sup>d</sup> (0.0) | 0.4 (0.0-1.2) | 0.3 (0.0-0.8) |
|  | EUR | 0.4 (1.4) | 0.3 (1.1) | 0.2 (0.8) | 0.2 (0.6) | 0.2 (0.5) | 0.1 (0.4) | 0.1 (0.3) | 1.5 (0.9-1.9) | 0.7 (0.4-0.8) |
|  | SEAR | 0.9 (1.0) | 0.5 (0.6) | 0.3 (0.4) | 0.2 (0.2) | 0.1 (0.1) | 0.1 (0.1) | <0.1<sup>c</sup> (0.1) | 2.1 (0.2-3.9) | 0.4 (0.0-0.8) |
|  | WPR | 0.7 (1.3) | 0.5 (0.9) | 0.4 (0.6) | 0.3 (0.4) | 0.2 (0.2) | 0.1 (0.2) | 0.1 (0.1) | 2.3 (1.3-3.3) | 0.5 (0.3-0.7) |
|  | Overall | 2.7 (0.9) | 1.9 (0.6) | 1.3 (0.5) | 1.0 (0.3) | 0.7 (0.3) | 0.5 (0.2) | 0.3 (0.1) | 8.4 (5.1-11.5) | 0.5 (0.3-0.6) |
| Males | AFR | <0.1<sup>c</sup> (0.1) | <0.1<sup>d</sup> (0.0) | <0.1<sup>d</sup> (0.0) | 0.0 (0.0) | 0.0 (0.0) | 0.0 (0.0) | 0.0 (0.0) | 0.0 (0.0-0.5) | 0.0 (0.0-0.3) |
|  | AMR | 0.5 (1.3) | 0.4 (1.0) | 0.3 (0.8) | 0.3 (0.7) | 0.2 (0.6) | 0.2 (0.5) | 0.1 (0.4) | 2.0 (1.3-2.4) | 0.8 (0.5-1.0) |
|  | EMR | 0.2 (0.7) | 0.1 (0.3) | 0.1 (0.2) | <0.1<sup>c</sup> (0.1) | <0.1<sup>c</sup> (0.0) | <0.1<sup>d</sup> (0.0) | <0.1<sup>d</sup> (0.0) | 0.4 (0.0-1.3) | 0.3 (0.0-0.8) |
|  | EUR | 0.3 (1.2) | 0.2 (0.9) | 0.2 (0.6) | 0.2 (0.5) | 0.1 (0.4) | 0.1 (0.3) | 0.1 (0.2) | 1.2 (0.7-1.5) | 0.5 (0.3-0.7) |
|  | SEAR | 1.0 (1.0) | 0.6 (0.6) | 0.3 (0.4) | 0.2 (0.2) | 0.1 (0.1) | 0.1 (0.1) | <0.1<sup>c</sup> (0.1) | 2.3 (0.2-4.2) | 0.4 (0.0-0.8) |
|  | WPR | 0.8 (1.3) | 0.8 (0.9) | 0.4 (0.6) | 0.3 (0.4) | 0.2 (0.2) | 0.1 (0.2) | 0.1 (0.1) | 2.5 (1.4-3.5) | 0.5 (0.3-0.7) |
|  | Overall | 2.9 (0.9) | 1.9 (0.6) | 1.3 (0.4) | 1.0 (0.3) | 0.6 (0.2) | 0.4 (0.2) | 0.3 (0.1) | 8.4 (5.0-11.8) | 0.4 (0.3-0.6) |
| Both | Global | 5.6 (0.9) | 3.8 (0.6) | 2.7 (0.5) | 2.0 (0.3) | 1.3 (0.2) | 0.9 (0.2) | 0.6 (0.1) | 16.8 (10.6-22.4) | 0.3 (0.2-0.4) |

Abbreviations: AFR = African Region, AMR = Region of the Americas, EMR = Eastern Mediterranean Region, EUR = European Region, HSV = Herpes simplex virus, SEAR = South-East Asia region, UI = Uncertainty interval, WHO = World Health Organization, WPR = Western Pacific Region.
<sup>a</sup> 95% UI of the total number of infected people in millions.
<sup>b</sup> 95% UI of percentage incidence.
<sup>c</sup> Numbers are <50,000 but ≥10,000.
<sup>d</sup> Numbers are <5,000 but ≥1,000.
Numbers (N) are the estimated number of people newly infected with HSV-1 during 2020. Numbers do not always sum exactly to the totals due to rounding.
Incidences (%) are the percentage of infected people in the age-, sex-, and region-specific population.
Regions are per World Health Organization definitions.

The number of people with prevalent genital HSV-1 infections globally in 2020 among individuals aged 15-49 years was estimated at 376.2 million (95% UI: 235.6-483.5 million), a genital HSV-1 prevalence of 10.2% (95% UI: 6.4-13.1%) (Table 4). Globally, genital HSV-1 prevalence was slightly higher in females (10.5%; 95% UI: 6.4-13.8%) than in males (9.9%; 95% UI: 5.9-13.4%). The Americas region had the highest genital HSV-1 prevalence, but the Western Pacific region had the largest number of infected persons among those aged 15-49 years.

**Table 4.** Global and regional estimates of the number and percentage of the population aged 15-49 years with prevalent genital HSV-1 infection in 2020, by age and sex.

|  |  | Number of people with prevalent genital HSV-1 infection in millions (population prevalence, %) by age group |  |  |  |  |  |  |  |  |
| --- | --- | --- | --- | --- | --- | --- | --- | --- | --- | --- |
| Sex | WHO region | 15-19 years | 20-24 years | 25-29 years | 30-34 years | 35-39 years | 40-44 years | 45-49 years | All ages |  |
|  |  | N (%) | N (%) | N (%) | N (%) | N (%) | N (%) | N (%) | N (95% UI<sup>a</sup>) | % (95% UI<sup>b</sup>) |
| Females | AFR | 0.1 (0.2) | 0.1 (0.2) | 0.1 (0.2) | 0.1 (0.3) | 0.1 (0.3) | 0.1 (0.3) | 0.1 (0.3) | 0.6 (0.0-9.4) | 0.3 (0.0-4.4) |
|  | AMR | 1.0 (3.5) | 3.5 (9.6) | 5.6 (14.6) | 7.0 (18.6) | 7.9 (22.0) | 8.3 (24.7) | 8.5 (26.9) | 41.7 (27.5-50.6) | 17.0 (11.2-20.6) |
|  | EMR | 0.5 (1.9) | 1.2 (4.3) | 1.6 (5.4) | 1.7 (5.9) | 1.5 (6.2) | 1.3 (6.3) | 1.1 (6.4) | 8.8 (0.4-22.5) | 5.7 (0.3-14.5) |
|  | EUR | 0.7 (3.8) | 2.4 (9.9) | 4.1 (14.5) | 5.9 (18.1) | 6.8 (20.8) | 7.3 (22.8) | 7.7 (24.4) | 35.1 (22.8-43.0) | 15.9 (10.3-19.5) |
|  | SEAR | 1.8 (2.7) | 5.1 (6.5) | 6.9 (8.8) | 7.8 (10.2) | 8.1 (11.1) | 7.7 (11.7) | 7.2 (12.0) | 44.7 (4.4-74.2) | 9.3 (0.9-15.4) |
|  | WPR | 1.5 (3.6) | 4.8 (8.8) | 7.6 (12.2) | 11.3 (14.4) | 10.4 (15.9) | 10.6 (16.9) | 13.1 (17.5) | 59.3 (35.1-80.1) | 12.0 (7.1-16.2) |
|  | Overall | 5.7 (1.9) | 17.2 (6.0) | 25.9 (9.0) | 33.8 (11.5) | 34.9 (13.1) | 35.3 (14.5) | 37.6 (15.9) | 190.3 (115.2-249.7) | 10.5 (6.4-13.8) |
| Males | AFR | 0.1 (0.2) | 0.1 (0.2) | 0.1 (0.2) | 0.1 (0.3) | 0.1 (0.3) | 0.1 (0.3) | 0.1 (0.3) | 0.6 (0.0-9.2) | 0.3 (0.0-4.3) |
|  | AMR | 1.0 (3.3) | 3.3 (8.9) | 5.2 (13.5) | 6.5 (17.2) | 7.2 (20.3) | 7.4 (22.8) | 7.4 (24.8) | 38.0 (24.8-45.8) | 15.6 (10.2-18.9) |
|  | EMR | 0.5 (1.9) | 1.3 (4.3) | 1.7 (5.4) | 1.8 (5.9) | 1.7 (6.2) | 1.4 (6.3) | 1.2 (6.4) | 9.7 (0.6-25.0) | 5.8 (0.3-14.9) |
|  | EUR | 0.6 (3.0) | 2.0 (7.9) | 3.4 (11.5) | 4.7 (14.2) | 5.3 (16.1) | 5.6 (17.6) | 5.8 (18.7) | 27.5 (16.8-34.0) | 12.4 (7.5-15.3) |
|  | SEAR | 2.0 (2.7) | 5.6 (6.5) | 7.5 (8.8) | 8.3 (10.2) | 8.5 (11.1) | 8.0 (11.7) | 7.3 (12.0) | 47.3 (3.7-78.6) | 9.3 (0.7-15.5) |
|  | WPR | 1.7 (3.6) | 5.3 (8.8) | 8.3 (12.2) | 12.0 (14.4) | 10.9 (15.9) | 11.0 (16.9) | 13.6 (17.5) | 62.9 (38.4-84.3) | 12.0 (7.3-16.1) |
|  | Overall | 5.9 (1.9) | 17.7 (5.8) | 26.3 (8.6) | 33.4 (10.9) | 33.7 (12.3) | 33.5 (13.5) | 35.4 (14.8) | 186.0 (111.4-250.7) | 9.9 (5.9-13.4) |
| Both | Global | 11.6 (1.9) | 34.9 (5.9) | 52.2 (8.8) | 67.2 (11.2) | 68.6 (12.7) | 68.8 (14.0) | 73.0 (15.3) | 376.2 (235.6-483.5) | 10.2 (6.4-13.1) |

Abbreviations: AFR = African Region, AMR = Region of the Americas, EMR = Eastern Mediterranean Region, EUR = European Region, HSV = Herpes simplex virus, SEAR = South-East Asia region, UI = Uncertainty interval, WHO = World Health Organization, WPR = Western Pacific Region.
<sup>a</sup> 95% UI of the total number of infected people in millions.
<sup>b</sup> 95% UI of percentage prevalence.
Numbers (N) are the year 2020 estimated number of people living with HSV-1 infection. Numbers do not always sum exactly to the totals due to rounding.
Prevalences (%) are the percentage of infected people in the age-, sex-, and region-specific population.
Regions are per World Health Organization definitions.

### All genital HSV infections

The number of incident genital HSV infections (HSV-2 and/or HSV-1) globally in 2020 among individuals aged 15-49 years was estimated at 42.4 million (95% UI: 33.7-51.8 million). The number of people with prevalent genital HSV infections (HSV-2 and/or HSV-1) globally in 2020 among individuals aged 15-49 years was estimated at 846.1 million (95% UI: 661.1-1034.2 million).

### HSV-2 and HSV-1 GUD

The number of individuals 15 to 49 years of age with at least one HSV-2 GUD episode in 2020 was estimated at 187.9 million (95% UI: 116.0-291.8 million), a prevalence of 4.8% (95% UI: 3.0-7.5%) (Table 5). Globally, HSV-2 GUD prevalence was considerably higher in females (6.2; 95% UI: 3.8-9.6%) than in males (3.5%; 95% UI: 2.2-5.8%). The total sum of GUD person-days was estimated at 8,675 million (95% UI: 5,632-15,068 million) (Table S7). The African region had both the highest HSV-2 GUD prevalence and the largest number of HSV-2 GUD person-days.

**Table 5.** Global and regional estimates of number and percentage of the population aged 15-49 years with at least one episode of GUD due to HSV-2 or HSV-1 in 2020, by age and sex.

| Sex | WHO region | Number of people with HSV GUD in millions (population prevalence, %) by age group |  |  |  |  |  |  |  |  |
| --- | --- | --- | --- | --- | --- | --- | --- | --- | --- | --- |
|  |  | 15-19 years | 20-24 years | 25-29 years | 30-34 years | 35-39 years | 40-44 years | 45-49 years | All ages |  |
|  |  | N (%) | N (%) | N (%) | N (%) | N (%) | N (%) | N (%) | N (95% UI<sup>a</sup>) | % (95% UI<sup>b</sup>) |
| HSV-2 |  |  |  |  |  |  |  |  |  |  |
| Females | AFR | 4.0 (6.8) | 6.0 (11.9) | 7.0 (16.0) | 7.3 (19.4) | 7.1 (22.2) | 6.4 (24.4) | 5.6 (26.3) | 43.3 (26.3-67.4) | 16.1 (9.8-25.0) |
|  | AMR | 1.0 (2.6) | 1.8 (4.7) | 2.7 (6.7) | 3.3 (8.6) | 3.8 (10.4) | 4.1 (12.0) | 4.3 (13.5) | 20.9 (12.2-33.9) | 8.2 (4.7-13.2) |
|  | EMR | 0.1 (0.3) | 0.2 (0.6) | 0.3 (0.9) | 0.3 (1.2) | 0.4 (1.5) | 0.4 (1.8) | 0.3 (2.1) | 2.0 (0.5-7.6) | 1.1 (0.3-4.1) |
|  | EUR | 0.3 (1.2) | 0.6 (2.2) | 0.9 (3.2) | 1.4 (4.1) | 1.7 (5.0) | 1.9 (5.9) | 2.2 (6.8) | 8.9 (4.5-16.6) | 4.2 (2.1-7.9) |
|  | SEAR | 1.0 (1.1) | 1.7 (2.1) | 2.4 (3.0) | 3.0 (3.9) | 3.5 (4.8) | 3.7 (5.6) | 3.9 (6.4) | 19.2 (11.2-32.0) | 3.6 (2.1-6.0) |
|  | WPR | 0.8 (1.4) | 1.5 (2.6) | 2.5 (3.8) | 3.9 (5.0) | 4.0 (6.0) | 4.5 (7.1) | 6.1 (8.1) | 23.3 (10.6-46.6) | 5.1 (2.3-10.1) |
|  | Overall | 7.1 (2.4) | 11.8 (4.1) | 15.7 (5.5) | 19.3 (6.5) | 20.4 (7.7) | 21.0 (8.6) | 22.3 (9.4) | 117.6 (72.0-184.1) | 6.2 (3.8-9.6) |
| Males | AFR | 2.3 (3.8) | 3.5 (6.9) | 4.2 (9.7) | 4.6 (12.2) | 4.6 (14.4) | 4.2 (16.5) | 3.8 (18.3) | 27.1 (15.9-43.1) | 10.1 (5.9-16.0) |
|  | AMR | 0.5 (1.3) | 1.0 (2.5) | 1.4 (3.6) | 1.8 (4.7) | 2.1 (5.7) | 2.2 (6.7) | 2.3 (7.7) | 11.3 (6.2-19.0) | 4.4 (2.4-7.4) |
|  | EMR | 0.1 (0.4) | 0.2 (0.7) | 0.3 (1.1) | 0.4 (1.4) | 0.5 (1.7) | 0.5 (2.1) | 0.4 (2.4) | 2.5 (1.0-6.3) | 1.3 (0.5-3.2) |
|  | EUR | 0.2 (0.6) | 0.3 (1.2) | 0.5 (1.8) | 0.8 (2.3) | 0.9 (2.8) | 1.1 (3.4) | 1.2 (3.9) | 5.0 (2.3-10.2) | 2.3 (1.0-4.7) |
|  | SEAR | 0.6 (0.6) | 1.0 (1.1) | 1.4 (1.6) | 1.7 (2.1) | 2.0 (2.6) | 2.1 (3.1) | 2.2 (3.6) | 11.1 (5.5-20.2) | 2.0 (1.0-3.6) |
|  | WPR | 0.5 (0.7) | 0.9 (1.4) | 1.4 (2.0) | 2.2 (2.6) | 2.2 (3.2) | 2.5 (3.8) | 3.4 (4.4) | 13.2 (4.4-36.2) | 2.7 (0.9-7.4) |
|  | Overall | 4.1 (1.3) | 7.0 (2.3) | 9.4 (3.1) | 11.5 (3.7) | 12.3 (4.5) | 12.6 (5.1) | 13.4 (5.6) | 70.3 (43.0-116.7) | 3.5 (2.2-5.8) |
| Both | Global | 11.2 (1.8) | 18.8 (3.2) | 25.1 (4.2) | 30.8 (5.1) | 32.7 (6.0) | 33.6 (6.8) | 35.7 (7.5) | 187.9 (116.0-291.8) | 4.8 (3.0-7.5) |
| HSV-1 |  |  |  |  |  |  |  |  |  |  |
| Females | AFR | 0.0 (0.0) | 0.0 (0.0) | 0.0 (0.0) | 0.0 (0.0) | 0.0 (0.0) | 0.0 (0.0) | 0.0 (0.0) | 0.0 (0.0-0.5) | 0.0 (0.0-0.3) |
|  | AMR | 0.4 (0.9) | 0.5 (1.2) | 0.4 (1.0) | 0.3 (0.8) | 0.2 (0.7) | 0.2 (0.5) | 0.1 (0.4) | 2.1 (1.2-3.0) | 0.8 (0.5-1.2) |
|  | EMR | 0.2 (0.5) | 0.1 (0.5) | 0.1 (0.2) | 0.0 (0.1) | 0.0 (0.1) | 0.0 (0.0) | 0.0 (0.0) | 0.4 (0.0-1.2) | 0.3 (0.0-0.8) |
|  | EUR | 0.3 (1.0) | 0.3 (1.2) | 0.3 (0.9) | 0.2 (0.7) | 0.2 (0.5) | 0.1 (0.4) | 0.1 (0.3) | 1.5 (0.8-2.2) | 0.7 (0.4-1.0) |
|  | SEAR | 0.6 (0.7) | 0.6 (0.7) | 0.4 (0.5) | 0.2 (0.3) | 0.1 (0.2) | 0.1 (0.1) | 0.0 (0.1) | 2.1 (0.2-4.1) | 0.4 (0.0-0.8) |
|  | WPR | 0.5 (0.9) | 0.6 (1.0) | 0.4 (0.7) | 0.4 (0.4) | 0.2 (0.3) | 0.1 (0.2) | 0.1 (0.1) | 2.3 (1.2-3.6) | 0.5 (0.2-0.7) |
|  | Overall | 1.9 (0.7) | 2.1 (0.7) | 1.5 (0.5) | 1.1 (0.4) | 0.8 (0.3) | 0.5 (0.2) | 0.4 (0.2) | 8.4 (4.7-12.5) | 0.5 (0.3-0.7) |
| Males | AFR | 0.0 (0.0) | 0.0 (0.0) | 0.0 (0.0) | 0.0 (0.0) | 0.0 (0.0) | 0.0 (0.0) | 0.0 (0.0) | 0.0 (0.0-0.6) | 0.0 (0.0-0.3) |
|  | AMR | 0.3 (0.9) | 0.4 (1.1) | 0.4 (0.9) | 0.3 (0.7) | 0.2 (0.6) | 0.2 (0.5) | 0.1 (0.4) | 1.9 (1.1-2.8) | 0.8 (0.5-1.1) |
|  | EMR | 0.2 (0.5) | 0.1 (0.5) | 0.1 (0.2) | 0.0 (0.1) | 0.0 (0.1) | 0.0 (0.0) | 0.0 (0.0) | 0.4 (0.0-1.4) | 0.3 (0.0-0.8) |
|  | EUR | 0.2 (0.8) | 0.3 (1.0) | 0.2 (0.7) | 0.2 (0.5) | 0.1 (0.4) | 0.1 (0.3) | 0.1 (0.2) | 1.2 (0.6-1.7) | 0.5 (0.3-0.8) |
|  | SEAR | 0.7 (0.7) | 0.7 (0.7) | 0.4 (0.5) | 0.2 (0.3) | 0.1 (0.2) | 0.1 (0.1) | 0.0 (0.1) | 2.2 (0.2-4.4) | 0.4 (0.0-0.9) |
|  | WPR | 0.6 (0.9) | 0.7 (1.0) | 0.5 (0.7) | 0.4 (0.4) | 0.2 (0.3) | 0.1 (0.2) | 0.1 (0.1) | 2.5 (1.3-4.0) | 0.5 (0.2-0.8) |
|  | Overall | 2.0 (0.6) | 2.2 (0.7) | 1.5 (0.5) | 1.1 (0.4) | 0.7 (0.3) | 0.5 (0.2) | 0.3 (0.1) | 8.3 (4.5-12.7) | 0.4 (0.2-0.7) |
| Both | Global | 3.9 (0.6) | 4.3 (0.7) | 3.1 (0.5) | 2.2 (0.4) | 1.4 (0.3) | 1.0 (0.2) | 0.7 (0.1) | 16.7 (9.3-25.2) | 0.5 (0.3-0.7) |
| HSV-2 and HSV-1 |  |  |  |  |  |  |  |  |  |  |
| Overall GUD | Females | 9.0 (3.1) | 14.0 (4.9) | 17.2 (6.0) | 20.4 (6.9) | 21.2 (8.0) | 21.5 (8.8) | 22.7 (9.6) | 126.0 (80.3-191.4) | 6.6 (4.2-10.1) |
|  | Males | 6.1 (1.9) | 9.2 (3.0) | 10.9 (3.6) | 12.6 (4.1) | 13.0 (4.7) | 13.1 (5.3) | 13.7 (5.7) | 78.6 (50.2-125.2) | 4.0 (2.5-6.3) |
|  | Both | 15.2 (2.5) | 23.2 (3.9) | 28.1 (4.8) | 33.0 (5.5) | 34.2 (6.3) | 34.5 (7.0) | 36.4 (7.6) | 204.6 (132.3-306.5) | 5.3 (3.4-7.9) |

Abbreviations: AFR = African Region, AMR = Region of the Americas, EMR = Eastern Mediterranean Region, EUR = European Region, GUD = Genital ulcer disease, HSV = Herpes simplex virus, SEAR = South-East Asia region, UI = Uncertainty interval, WHO = World Health Organization, WPR = Western Pacific Region.
<sup>a</sup> 95% UI of the total number of infected people in millions.
<sup>b</sup> 95% UI of percentage prevalence.
Numbers (N) are the year 2020 estimated number of people living with genital ulcer disease caused by HSV-1 and/or HSV-2. Numbers do not always sum exactly to the totals due to rounding.
Prevalences (%) are the percentage of infected people in the age-, sex-, and region-specific population.
Regions are per World Health Organization definitions.

The number of individuals 15 to 49 years of age with at least one HSV-1 GUD episode in 2020 was estimated at 16.7 million (95% UI: 9.3-25.2 million), a prevalence of 0.5% (95% UI: 0.3-0.7%) (Table 5). The total sum of GUD person-days was estimated at 166 million (95% UI: 98-562 million) (Table S7). The Americas region had the highest HSV-1 GUD prevalence and the Western Pacific region had the largest number of HSV-1 GUD person-days.

The number of individuals 15 to 49 years of age with at least one HSV-2 or HSV-1 GUD episode in 2020 was estimated to be 204.6 (95% UI: 132.3-306.5) million (Table 5). The total number of person-days with GUD was estimated at 8,841 (95% UI: 5,795-15,425) (Table S7).

### All and only oral HSV-1 infections

Number of new (incident) HSV-1 infections globally in 2020 at any site (oral and genital) among individuals aged 0-49 years was estimated at 122.2 million (95% UI: 116.2-128.6 million) (Table S8). The African region had the highest HSV-1 incidence at nearly 40 million infections. HSV-1 incidence rate decreased with age, most notably in regions where prevalence reached saturation at younger ages such as the African and Eastern Mediterranean regions (Table S8 and Figure S1).

The number of people with prevalent HSV-1 infections globally in 2020 at any site (oral and genital) among individuals aged 0-49 years was estimated at 3,779.1 million (95% UI: 3,510.3-3,921.6 million), a prevalence of 64.2% (95% UI: 59.7-66.7%) (Table S9). The African had the highest prevalence, but the Western Pacific region had the largest number of HSV-1 infected persons. In the secondary analysis among individuals aged 50-99 years, an additional 1,523.6 million people were estimated to be infected worldwide (Table S6).

Global prevalence of oral HSV-1 infection in 2020 among individuals aged 0-49 years was estimated at 58.6% (95% UI: 53.5-62.1%) (Table S10). A total of 3,448.9 million (95% UI: 3,144.9-3,655.2 million) people aged 0-49 years were estimated to be orally infected worldwide. The African region had the highest oral HSV-1 prevalence.

## Discussion

In 2020, we estimated that 26 million people aged 15 to 49 years acquired a new HSV-2 infection, 520 million people were living with an HSV-2 infection, and 188 million people had at least one episode of GUD caused by HSV-2. Additionally, in 2020, 17 million people aged 15 to 49 years acquired a new genital HSV-1 infection through sexual transmission, 376 million people were living with genital HSV-1 infection, and 17 million people had at least one episode of GUD caused by HSV-1. Notably, sexual transmission accounted for only a portion of all HSV-1 infections. In total, two-thirds of the global population aged 0-49 years, or nearly 4 billion people, were infected (mostly orally) with HSV-1 in 2020, with over 120 million individuals newly infected in this year.

The estimates for genital infection, and particularly for GUD, are higher for HSV-2 than for HSV-1. Almost all HSV-2 acquisitions are sexually transmitted and occur genitally, while the majority of HSV-1 acquisitions are not genitally acquired. Most HSV-1 infections are acquired orally in childhood, although an increasing number are being acquired through sexual activity in adolescence and adulthood [26–28, 35]. In addition, HSV-1 genital tract infection is much less likely to recur compared to HSV-2 infection [4], further reducing the relative contribution of HSV-1 to GUD, even in the presence of numerous genital HSV-1 infections.

HSV-2 global prevalence was virtually equal in the 2016 and 2020 estimation rounds (Table S5). Considering the shifts in the underlying demography during this time (increase in global average age and changing proportion of the global population in each region [47]), HSV-2 prevalence, adjusted for the demographic trends, appears to be slowly declining, as indicated also by meta-regression and modeling analyses applied on prevalence data [10-12, 34, 49]. This decline may reflect less risky sexual behaviour following the HIV epidemic [50–53], improved STI awareness [54], increasing access to HIV/STI services [55, 56], and/or changes in the structure of sexual networks following changes in socio-economic conditions [10].

The estimated number of prevalent genital HSV-1 infections is nearly two-fold higher in 2020 compared to 2016 (376 versus 192 million) (Table S5). Meta-regression analyses on the data extracted through the regional systematic reviews as well as modeling analyses have also indicated increasing rates of genital HSV-1 infection and decreasing rates of oral infections in several regions [26–28, 35], suggesting an epidemiological transition for this infection from an oral to increasingly genital acquisition [26, 57, 58]. The increase in the global adult population and its average age [47] has also contributed to the higher number of prevalent genital HSV-1 cases.

Differences in the global or regional estimates between 2016 and 2020 for both infections may also be attributed to improved data availability, specifically for capturing HSV-1 genital infections. In 2020, we had considerably more data from children in the South-East Asia region and the Western Pacific region, potentially presenting a more realistic pattern of HSV-1 prevalence by age in these regions. Although there was some overlap in the data used for the 2016 and 2020 rounds, these inputs were sourced from different countries and diverse general populations. Caution is warranted in interpreting differences or similarity in time trends of all estimates considering the changes in input data.

This study has several limitations. The primary challenge in estimating HSV-1 and HSV-2 prevalence and incidence lies in the availability and representativeness of the data. However, the enhancement in the 2020 round of calibrating the model based on the series of standardized and comprehensive HSV-1 and HSV-2 systematic reviews covering all global regions [9-12, 27, 28, 34-40] has improved on this limitation with more data available than was used in previous rounds (although this was also partly due to an expanded time frame for including data in the 2020 estimates). The challenges associated with input data underscore the need for more substantial, high quality, and representative epidemiological data pertaining to HSV infections.

Some of the estimates, particularly those relating to HSV-1 genital infection, had wide 95% UIs due to limited input data. HSV-1 prevalence data among children remain very limited, but these data are critical for accurate and precise estimates of HSV-1 genital infection rates. Adults can be infected genitally only if they were not infected orally during childhood [26]. The proportion of incident HSV-1 infections that were estimated to be genital versus oral in adulthood was based on pooled data from only four available longitudinal studies, all of which were from the United States of America and based only on symptomatic infection incidence [1, 59–62]. The estimate for this proportion may not be representative of its value in other countries or of asymptomatic infection.

The force of infection was assumed constant by age, but the force of infection was only applied to those still susceptible to infection thereby effectively allowing incidence rate to decrease with age. The model was also calibrated for the maximum proportion that can be infected in addition to the force of infection. This ensured that prevalence saturates below 100% where indicated by prevalence data.

The model was designed to provide estimates only for those aged under 50 years, as this age group is the focus of WHO programs for sexual transmission of infection and reproductive health outcomes, and to align HSV estimates with WHO estimates for other STIs [63–65]. Prevalence estimates were additionally calculated in a secondary exploratory analysis for those aged above 50 years. However, these estimates were based on simplifying assumptions due to very limited prevalence data for those aged above 50 years. No incidence estimates could be generated, and no uncertainty bounds were incorporated for these prevalence estimates due to the simplifying assumptions in generating them.

In conclusion, HSV infections are widely prevalent in all global regions, leading to a significant burden of GUD with repercussions on psychosocial, sexual, and reproductive health, neonatal transmission, and HIV transmission. However, hardly any specific programs for HSV prevention and control exist, even in resource-rich countries [66, 67], partly due to the lack of tools to address such highly prevalent, often asymptomatic, and incurable infections on a population level. Available prevention modalities, including condoms and antiviral therapy, are insufficient to control infection transmission and have, at best, had a modest population impact in reducing incidence rates. There is a critical need for HSV prophylactic and therapeutic vaccines as a strategic approach to control transmission and to curb the disease and economic burdens of these infections [68–70].

## List of abbreviations

CI: Confidence intervals
GATHER: Guidelines for Accurate and Transparent Health Estimates Reporting
GUD: Genital ulcer disease
HSV: Herpes simplex virus
HSV-1: Herpes simplex virus type 1
HSV-2: Herpes simplex virus type 2
UI: Uncertainty interval
WHO: World Health Organization

## Supplementary Information

**Ad**ditional file 1: Supplementary Material

## Acknowledgements

The authors gratefully acknowledge Professor Emeritus Rhoda Ashley Morrow from the University of Washington, for her support in assessing the quality of study diagnostic methods, and Dr. Maeve Brito De Mell from the World Health Organization for her invaluable insights during the planning and implementation of this project. The authors are also grateful to Ms. Adona Canlas for administrative support. This publication was supported by the World Health Organization (Global HIV, Hepatitis and STIs Programmes) through a grant from USAID. WHO commissioned the study, advised as required, helped with redrafts, and approved manuscript submission. MH, SM, AA, AMMO, and LJA are grateful for research support provided by the Biomedical Research Program and by the Biostatistics, Epidemiology, and the Biomathematics Research Core at Weill Cornell Medicine-Qatar. KJL thanks the National Institute for Health Research (NIHR) and the Health Protection Research Unit (HPRU) in Behavioural Science and Evaluation at the University of Bristol, in partnership with the UK Health Security Agency (UKHSA), for research support. The original systematic reviews for HSV-1 and HSV-2 were funded in part by grant numbers ARG01-0524-230321 from the Qatar Research, Development, and Innovation Council and NPRP9-040-3-008 from the Qatar National Research Fund (a member of Qatar Foundation). The authors alone are responsible for the views expressed in this article and they do not necessarily represent the views, decisions or policies of the institutions with which they are affiliated, including the NHS, the NIHR, the Department for Health and Social Care, the UKHSA, and the WHO.

## Data Availability Statement

This mathematical modeling study is based on data that are publicly available. Data is provided within the manuscript or supplementary information files. No Institutional Review Board clearance was required to use these data.

## Authors’ Contributions

MH, SM, AA, and AMMO conducted the literature searches, data extraction, data synthesis, and meta-analyses. KJL conducted the stages of estimates calculation and contributed to writing of the first draft. MH and LJA wrote the first draft of the manuscript. SLG and JR provided input and guidance at all stages. All authors contributed to discussion and interpretation of the results, editing of the article, and have read and approved the final manuscript.

## Ethics approval and consent to participate

Not applicable.

## Consent for publication

Not applicable.

## Supplementary Material

### Box S1.

Summary of the standardized methodology used in updating the previously conducted systematic reviews for Asia [1, 2], Europe [3, 4], Latin America and the Caribbean [5, 6], Middle East and North Africa [7, 8], sub-Saharan Africa [9, 10], and for Canada, Australia, New Zealand, and Pacific Island nations [11–13].

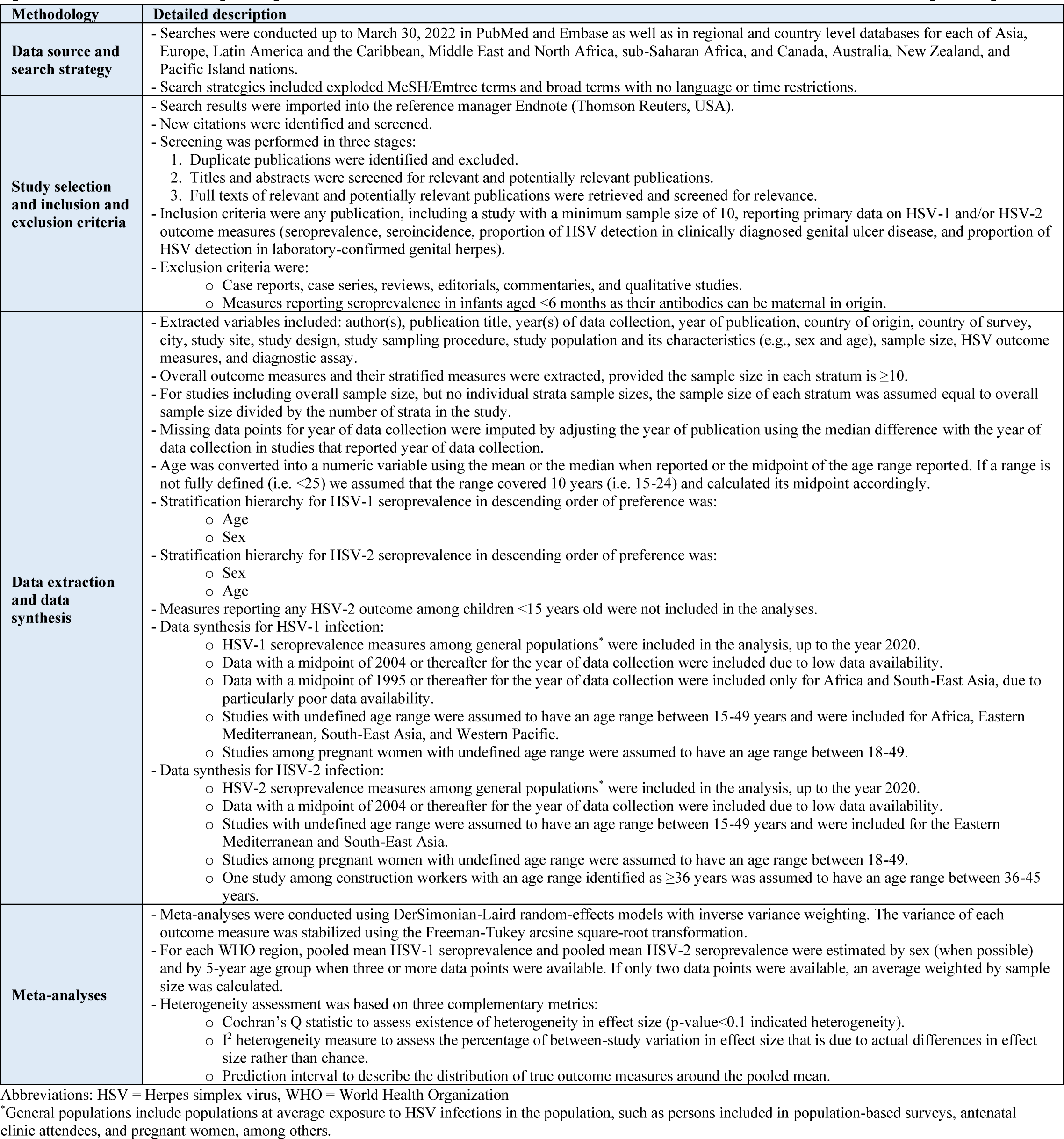

### Box S2

List of changes and methodological enhancements on the 2016 WHO HSV estimation round [15].

#### Preparation of the seroprevalence data for pooling

1. Equivocal samples were included in the denominator of seroprevalence data points, rather than being excluded from both numerator and denominator, to avoid bias as such equivocal samples are likely to be sero-negative.
2. Adjustment for sensitivity and specificity for each assay type was not conducted since only studies using a robust assay were included. Moreover, assay type showed no association with HSV seroprevalence based on the series of conducted regional systematic reviews and meta-regressions for HSV-1 and HSV-2 seroprevalence [1–13].
3. Meta-analyses were conducted when three or more data points were available. Average seroprevalence (weighted by sample size) was calculated when only two data points were available.
4. Missing data points for year of data collection were imputed by adjusting the year of publication using the median difference with the year of data collection in studies that reported year of data collection.
5. For studies of pregnant women where data on age were not available, we assumed an age range of 18-49 years.

#### Decisions on pooling criteria

##### HSV-1 estimates

1. Data with a midpoint of 2004 or thereafter for the year of data collection were included due to low data availability.
2. Data with a midpoint of 1995 or thereafter for the year of data collection were included only for Africa and South-East Asia, due to particularly poor data availability.
3. Studies with undefined age range were assumed to have an age range between 15-49 years and were included for Africa, Eastern Mediterranean, South-East Asia, and Western Pacific.
4. Single data points were included in the fitting for South-East Asia, due to particularly poor data availability.
5. Sex-specific estimates were produced for the Americas and Europe as per previous estimation round (with mixed-sex estimates for the other regions), except for children (<15 years), where mixed-sex pooled mean estimates were calculated for a given age band. These combined estimates were used in the fitting for each of females and males, to avoid the few data points for children by sex skewing the model fits by sex.
6. No sex specific estimates were calculated for Africa, Eastern Mediterranean region, South-East Asia, and Western Pacific since previous systematic reviews and meta-regressions showed no difference in seroprevalence by sex for these regions [2, 7, 10, 13].

##### HSV-2 estimates

1. Data with a midpoint of 2004 or thereafter for the year of data collection were included due to low data availability.
2. Studies with undefined age range were assumed to have an age range between 15-49 years and were included for Eastern Mediterranean and South-East Asia.
3. Single data points were included in the fitting for Eastern Mediterranean, due to particularly poor data availability.

Abbreviations: HSV = Herpes simplex virus, WHO = World Health Organization

### Box S3

Calculation of prevalence and incidence estimates

Pooled mean herpes simplex virus (HSV) seroprevalence values were used to calibrate HSV-1 and HSV-2 incidence using a constant incidence rate model. This model additionally included a term representing the maximum proportion of individuals that can be infected which was simultaneously calibrated. This term allows seroprevalence to saturate at a low or moderate value if this is indicated by the pooled seroprevalence values, for example as a consequence of lower incidence at older age. The model equation was as follows:

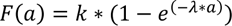

where *F(a)* is the proportion of individuals (sero)positive at age *a* in years (i.e., with prevalent HSV-1 or HSV-2 infection), *k* is the maximum proportion of individuals that can be infected and λ is the force of infection per year. We assumed that individuals can be infected with HSV-1 from age 0 years, and with HSV-2 from age 12 years, and adjusted the above equation as follows for HSV-2:

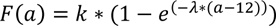

Fitting was done by using the Solver function in Excel to find those values of *k* and λ which maximized the (Bernoulli likelihood) value of:

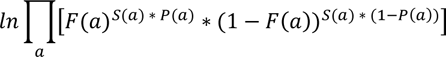

where *a* is the mid-point of each 5-year age group, *S(a*) is the total sample size (from summing across all studies), and *P(a)* is the pooled seroprevalence.

From the model fits we obtained smoothed seroprevalence estimates by sex and 5-year age group. Using demographic data for 2020 by sex and 5-year age group obtained from the United Nations Population Division [14], and assuming a uniform distribution for population size within each 5-year age group, we multiplied seroprevalence estimates at each single year of age by regional population sizes to obtain estimates for the number of people with prevalent HSV-1 and HSV-2 infection by World Health Organization (WHO) region in 2020. The estimated number of new (incident) cases of infection at each single year of age, *I(a)*, was calculated as:

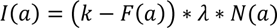

where *N(a)* is the total number of individuals (i.e., regional population size) at age *a*.

The percentage of people with genital infection due to either HSV-1 or HSV-2 was calculated using the following formula to correct for the estimated percentage with both HSV-2 and genital HSV-1 infection: % with any genital HSV infection = [(% with HSV-2 infection + % with genital HSV-1 infection)] – (% with HSV-2 infection * % with genital HSV-1 infection)

Calculations were done for each single year of age separately by WHO region and sex. These percentages were then applied to demographic data for 2020 [14] and summed.

#### Uncertainty bounds

We computed 95% uncertainty intervals (UI) on the number of individuals with prevalent HSV-1 and HSV-2 infection in 2020, as a function of the uncertainty in the underlying seroprevalence data. First, we recalibrated the force of infection, *λ*, to the upper confidence bounds of each region-, age-, and sex-specific pooled HSV-1 or HSV-2 seroprevalence value, keeping *k* constant. We then repeated this for the lower confidence bounds of each pooled seroprevalence value. Next, we sampled the log force of infection using the log fitted force of infection and the standard error of the log fitted force of infection by sex and WHO region, assuming a normal distribution. The proportion of individuals from age 15 years that are infected orally and the proportion that are infected genitally among those with incident HSV-1 infection were also simultaneously sampled, again using the log odds and standard error of the log odds from the results of the meta-analysis for these proportions [14]. Repeating this 1000 times, the resulting set of 1000 estimates was then sorted from low to high for each estimate, sex, and region of interest and the 2.5 and 97.5 percentiles taken to represent the lower and upper uncertainty bounds.

**Table S1.** Guidelines for Accurate and Transparent Health Estimates Reporting (GATHER) checklist.[16].

| Item # | Checklist item | Reported on page # |
| --- | --- | --- |
| **Objectives and funding** |  |  |
| 1 | Define the indicator(s), populations (including age, sex, and geographic entities), and time period(s) for which estimates were made. | Introduction section |
| 2 | List the funding sources for the work. | Title page |
| **Data Inputs** |  |  |
| For all data inputs from multiple sources that are synthesized as part of the study: |  |  |
| 3 | Describe how the data were identified and how the data were accessed. | Methods: Systematic reviews, data extraction and synthesis, and meta-analyses section; Box S1 |
| 4 | Specify the inclusion and exclusion criteria. Identify all ad-hoc exclusions. | Methods: Systematic reviews, data extraction and synthesis, and meta-analyses section; Box S1 |
| 5 | Provide information on all included data sources and their main characteristics. For each data source used, report reference information or contact name/institution, population represented, data collection method, year(s) of data collection, sex and age range, diagnostic criteria or measurement method, and sample size, as relevant. | Tables S1- S3; Box S4 and S5 |
| 6 | Identify and describe any categories of input data that have potentially important biases (e.g., based on characteristics listed in item 5). | Box S2 |
| For data inputs that contribute to the analysis but were not synthesized as part of the study: |  |  |
| 7 | Describe and give sources for any other data inputs. | Box S3 |
| For all data inputs: |  |  |
| 8 | Provide all data inputs in a file format from which data can be efficiently extracted (e.g., a spreadsheet rather than a PDF), including all relevant meta-data listed in item 5. For any data inputs that cannot be shared because of ethical or legal reasons, such as third-party ownership, provide a contact name or the name of the institution that retains the right to the data. | Tables S1- S3; Box S4 and S5 |
| **Data analysis** |  |  |
| 9 | Provide a conceptual overview of the data analysis method. A diagram may be helpful. | Methods: Population-based estimation for HSV-1 and HSV-2 infections section |
| 10 | Provide a detailed description of all steps of the analysis, including mathematical formulae. This description should cover, as relevant, data cleaning, data pre-processing, data adjustments and weighting of data sources, and mathematical or statistical model(s). | Box S3 |
| 11 | Describe how candidate models were evaluated and how the final model(s) were selected. | NA |
| 12 | Provide the results of an evaluation of model performance, if done, as well as the results of any relevant sensitivity analysis. | NA |
| 13 | Describe methods for calculating uncertainty of the estimates. State which sources of uncertainty were, and were not, accounted for in the uncertainty analysis. | Methods: Population-based estimation for HSV-1 and HSV-2 infections section; Box S3 |
| 14 | State how analytic or statistical source code used to generate estimates can be accessed. | Box S3 |
| **Results and Discussion** |  |  |
| 15 | Provide published estimates in a file format from which data can be efficiently extracted. | Tables 1-5; Tables S5-S7 |
| 16 | Report a quantitative measure of the uncertainty of the estimates (e.g. uncertainty intervals). | Tables 1-5; Tables S5-S7 |
| 17 | Interpret results in light of existing evidence. If updating a previous set of estimates, describe the reasons for changes in estimates. | Results section; Discussion section |
| 18 | Discuss limitations of the estimates. Include a discussion of any modelling assumptions or data limitations that affect interpretation of the estimates. | Discussion section |

Abbreviations: HSV = Herpes simplex virus, NA: Not Applicable.

**Table S2.** Summary of the extracted HSV-1 and HSV-2 epidemiological outcome measures in the series of regional systematic reviews and their updates.

| Region | Screened citations for HSV-1 and HSV-2 | Previously extracted relevant publications in the regional systematic reviews | Newly extracted relevant publications in the updates of the systematic reviews | Extracted epidemiological outcome measures |
| --- | --- | --- | --- | --- |
| Asia | 16,375 | 222 | 20 | • 26 overall HSV-2 seroincidence measures<br>• 66 overall HSV-1 seroprevalence measures yielding 204 stratified measures<br>• 283 overall HSV-2 seroprevalence measures yielding 697 stratified measures<br>• 9 overall proportions of HSV-1 detection in GUD cases<br>• 15 overall proportions of HSV-2 detection in GUD cases yielding 16 stratified proportions<br>• 26 overall proportions of HSV-1 detection in genital herpes cases yielding 27 stratified proportions<br>• 43 overall proportions of HSV-2 detection in genital herpes cases yielding 46 stratified proportions |
| Africa | 24,474 | 255 | 19 | • 95 overall HSV-2 seroincidence measures<br>• 49 overall HSV-1 seroprevalence measures yielding 76 stratified measures<br>• 352 overall HSV-2 seroprevalence measures yielding 811 stratified measures<br>• 19 overall proportions of HSV-1 detection in GUD cases yielding 32 stratified proportions<br>• 26 overall proportions of HSV-2 detection in GUD cases<br>• 8 overall proportions of HSV-1 detection in genital herpes cases<br>• 9 overall proportions of HSV-2 detection in genital herpes cases yielding 11 stratified proportions |
| Canada, Australia, New Zealand, and Pacific Islands | 3,173 | 90 | 55 | • 2 overall HSV-1 seroincidence measures<br>• 7 overall HSV-2 seroincidence measures<br>• 61 overall HSV-1 seroprevalence measures yielding 106 stratified seroprevalence measures<br>• 37 overall HSV-2 seroprevalence measures yielding 99 stratified seroprevalence measures<br>• 5 overall proportions of HSV-1 detection in GUD cases<br>• 5 overall proportions of HSV-2 detection in GUD cases<br>• 17 overall proportions of HSV-1 detection in genital herpes cases yielding 52 stratified proportions<br>• 16 overall proportions of HSV-2 detection in genital herpes cases yielding 46 stratified proportions |
| Europe | 19,153 | 353 | 15 | • 1 overall HSV-1 seroincidence measure<br>• 14 overall HSV-2 seroincidence measures<br>• 198 overall HSV-1 seroprevalence measures yielding 648 stratified measures<br>• 302 overall HSV-2 seroprevalence measures yielding 822 stratified measures<br>• 8 overall proportions of HSV-1 detection in GUD cases<br>• 17 overall proportions of HSV-2 detection in GUD cases yielding 19 stratified proportions<br>• 70 overall proportions of HSV-1 detection in genital herpes cases yielding 180 stratified proportions<br>• 76 overall proportions of HSV-2 detection in genital herpes cases yielding 201 stratified proportions |
| Middle East and North Africa | 5,320 | 52 | 8 | • 62 overall HSV-1 seroprevalence measures yielding 112 stratified measures<br>• 60 overall HSV-2 seroprevalence measures yielding 131 stratified measures<br>• 1 overall proportion of HSV-1 detection in GUD cases<br>• 1 overall proportion of HSV-2 detection in GUD cases<br>• 1 overall proportion of HSV-1 detection in genital herpes cases<br>• 3 overall proportions of HSV-2 detection in genital herpes cases yielding 6 stratified proportions |
| Latin America and the Caribbean | 8,477 | 133 | 6 | • 13 overall HSV-2 seroincidence measures<br>• 47 overall HSV-1 seroprevalence measures yielding 73 stratified measures<br>• 167 overall HSV-2 seroprevalence measures yielding 410 stratified measures<br>• 18 overall proportions of HSV-1 detection in GUD cases<br>• 7 overall proportions of HSV-2 detection in GUD cases<br>• 8 overall proportions of HSV-1 detection in genital herpes cases<br>• 10 overall proportion of HSV-2 detection in genital herpes cases |

Abbreviations: HSV = Herpes simplex virus, GUD = Genital ulcer disease.

### Box S4

List of articles on the general population from which an HSV-2 prevalence measure was extracted and included in the meta-analyses

1. Abbai NS, Govender S, Nyirenda M. Herpes simplex virus-2 infections in pregnant women from Durban, South Africa: prevalence, risk factors and co-infection with HIV-1. Southern African Journal of Infectious Diseases 2018.
2. Abdool Karim Q, Kharsany AB, Leask K, et al. Prevalence of HIV, HSV-2 and pregnancy among high school students in rural KwaZulu-Natal, South Africa: a bio-behavioural cross-sectional survey. Sex Transm Infect 2014; 90(8): 620-6.
3. Abul-Razak SHH, Abbas AA-D, Hwaid AH, Wasan AM, Fadeel ZG. Seroprevalence of Anti-Herpes Simplex Virus Type2 IgG, IgM Antibodies Among Pregnant Women in Diyala Province. Diyala Journal of Medicine 2013; 5(1): 36-43.
4. Achilles SL, Mhlanga F, Dezzutti CS, et al. Differences in genital tract immune cell populations and innate cervicovaginal fluid anti-HIV activity among women from Zimbabwe and the United States. AIDS Research and Human Retroviruses 2016; 32(Supplement 1): 78.
5. Adamson PC, Krupp K, Freeman AH, Klausner JD, Reingold AL, Madhivanan P. Prevalence & correlates of primary infertility among young women in Mysore, India. Indian Journal of Medical Research 2011; 134(10): 440-6.
6. Akinyi B, Odhiambo C, Otieno F, et al. Prevalence, incidence, and correlates of HSV-2 infection in an HIV incidence adolescent and adult cohort study in western Kenya. PloS one 2017; 12(6): e0178907.
7. Al-Hakami AM, Paul E, Al-Abed F, et al. Prevalence of toxoplasmosis, rubella, cytomegalovirus, and herpes (TORCH) infections among women attending the antenatal care clinic, maternity hospital in Abha, Southwestern Saudi Arabia. Saudi Med J 2020; 41(7): 757-62.
8. Alberts CJ, Schim van der Loeff MF, Papenfuss MR, et al. Association of Chlamydia trachomatis infection and herpes simplex virus type 2 serostatus with genital human papillomavirus infection in men: the HPV in men study. Sex Transm Dis 2013; 40(6): 508-15.
9. Ali MK, Hathal HD, Almoayed HA. Prevalence and diagnosis of genital herpes by immunological and molecular study Iraqi Journal of Medical Sciences 2018; 16(1): 4-7.
10. Alshareef SA, Eltom AM, Nasr AM, Hamdan HZ, Adam I. Rubella, herpes simplex virus type 2 and preeclampsia. Virol J 2017; 14(1): 142.
11. Anaedobe CG, Ajani TA. Co-infection of herpes simplex virus type 2 and HIV infections among pregnant women in Ibadan, Nigeria. Journal of Global Infectious Diseases 2019; 11(1): 19-24.
12. Anjulo AA, Abebe T, Hailemichael F, Mihret A. Seroprevalence and risk factors of herpes simplex virus-2 among pregnant women attending antenatal care at health facilities in Wolaita zone, Ethiopia. Virol J 2016; 13: 43.
13. Arama V, Cercel AS, Vladareanu R, et al. Type-specific herpes simplex virus-1 and herpes simplex virus-2 seroprevalence in Romania: comparison of prevalence and risk factors in women and men. International journal of infectious diseases: IJID: official publication of the International Society for Infectious Diseases 2010; 14 Suppl 3: e25-31.
14. Asgari S, Chamani-Tabriz L, Asadi S, et al. HSV-2 seroepidemiology and risk factors among Iranian women: A time to new thinking. Iranian Red Crescent Medical Journal 2011; 13(11): 818-23.
15. Austrian K, Hewett PC, Soler-Hampejsek E, Bozzani F, Behrman JR, Digitale J. Adolescent Girls Empowerment Programme: research and evaluation mid-term technical report, 2016.
16. Baird SJ, Garfein RS, McIntosh CT, Ozler B. Effect of a cash transfer programme for schooling on prevalence of HIV and herpes simplex type 2 in Malawi: a cluster randomised trial. Lancet (London, England) 2012; 379(9823): 1320-9.
17. Balaeva T, Grjibovski AM, Sidorenkov O, et al. Seroprevalence and correlates of herpes simplex virus type 2 infection among young adults in Arkhangelsk, Northwest Russia: a population-based cross-sectional study. 2016; 16(1): 616.
18. Behanzin L, Diabate S, Minani I, et al. Decline in HIV Prevalence among Young Men in the General Population of Cotonou, Benin, 1998-2008. PloS one 2012; 7(8): e43818.
19. Benjamin RJ, Busch MP, Fang CT, et al. Human immunodeficiency virus-1 infection correlates strongly with herpes simplex virus-2 (genital herpes) seropositivity in South African and United States blood donations. Transfusion 2008; 48(2): 295-303.
20. Biraro S, Kamali A, White R, et al. Effect of HSV-2 on population-level trends in HIV incidence in Uganda between 1990 and 2007. Tropical medicine & international health: TM & IH 2013; 18(10): 1257-66.
21. Birdthistle IJ, Floyd S, Machingura A, Mudziwapasi N, Gregson S, Glynn JR. From affected to infected? Orphanhood and HIV risk among female adolescents in urban Zimbabwe. Aids 2008; 22(6): 759-66.
22. Biswas D, Borkakoty B, Mahanta J, et al. Seroprevalence and risk factors of herpes simplex virus type-2 infection among pregnant women in Northeast India. BMC Infect Dis 2011; 11: 325.
23. Bjerke SE, Holter E, Vangen S, Stray-Pedersen B. Sexually transmitted infections among Pakistani pregnant women and their husbands in Norway. International journal of women’s health 2010; 2: 303-9.
24. Bochner AF, Madhivanan P, Niranjankumar B, et al. The Epidemiology of Herpes Simplex Virus Type-2 Infection among Pregnant Women in Rural Mysore Taluk, India. Journal of sexually transmitted diseases 2013; 2013: 750415.
25. Boni Cisse C, Zaba F, Meite S, et al. Seroprevalence of herpes simplex virus 2 infection among pregnant women in urban health training Yopougon-Attie (Cote D’ivoire). Academic Journals 2015; 6(3): 17-21.
26. Bradley J, Floyd S, Piwowar-Manning E, et al. Sexually transmitted bedfellows: exquisite association between HIV and herpes simplex virus type 2 in 21 communities in Southern Africa in the HIV prevention trials network 071 (PopART) Study. Journal of Infectious Diseases 2018; 218(3): 443-52.
27. Braunstein SL, Ingabire CM, Geubbels E, et al. High burden of prevalent and recently acquired HIV among female sex workers and female HIV voluntary testing center clients in Kigali, Rwanda. PloS one 2011a; 6(9): e24321.
28. Celentano DD, Mayer KH, Pequegnat W, et al. Prevalence of Sexually Transmitted Diseases and Risk Behaviors from the NIMH Collaborative HIV/STD Prevention Trial. International journal of sexual health: official journal of the World Association for Sexual Health 2010; 22(4): 272-84.
29. Centers for Disease Control and Prevention (CDC). National Center for Health Statistics (NCHS). National Health and Nutrition Examination Survey Data. Hyattsville, MD: U.S. Department of Health and Human Services, Centers for Disease Control and Prevention. 2008.
30. Centers for Disease Control and Prevention (CDC). National Center for Health Statistics (NCHS). National Health and Nutrition Examination Survey Data. Hyattsville, MD: U.S. Department of Health and Human Services, Centers for Disease Control and Prevention. 2010.
31. Centers for Disease Control and Prevention (CDC). National Center for Health Statistics (NCHS). National Health and Nutrition Examination Survey Data. Hyattsville, MD: U.S. Department of Health and Human Services, Centers for Disease Control and Prevention. 2012.
32. Centers for Disease Control and Prevention (CDC). National Center for Health Statistics (NCHS). National Health and Nutrition Examination Survey Data. Hyattsville, MD: U.S. Department of Health and Human Services, Centers for Disease Control and Prevention. 2014.
33. Centers for Disease Control and Prevention (CDC). National Center for Health Statistics (NCHS). National Health and Nutrition Examination Survey Data. Hyattsville, MD: U.S. Department of Health and Human Services, Centers for Disease Control and Prevention. 2016.
34. Chawla R, Bhalla P, Bhalla K, Singh MM, Garg S. Community-based study on seroprevalence of herpes simplex virus type 2 infection in New Delhi. Indian J Med Microbiol 2008; 26(1): 34-9.
35. Chen L, Liu J, Shi L, et al. Seasonal influence on TORCH infection and analysis of multi-positive samples with indirect immunofluorescence assay. J Clin Lab Anal 2019; 33(4): e22828.
36. Cheslack-Postava K, Brown AS, Chudal R, et al. Maternal exposure to sexually transmitted infections and schizophrenia among offspring. Schizophrenia research 2015; 166(1-3): 255-60.
37. Conde-Glez C, Lazcano-Ponce E, Rojas R, et al. Seroprevalences of varicella-zoster virus, herpes simplex virus and cytomegalovirus in a cross-sectional study in Mexico. Vaccine 2013; 31(44): 5067-74.
38. Crucitti T, Jespers V, Mulenga C, Khondowe S, Vandepitte J, Buve A. Non-sexual transmission of Trichomonas vaginalis in adolescent girls attending school in Ndola, Zambia. PloS one 2011; 6(1): e16310.
39. Dargham SR, Nasrallah GK, Al-Absi ES, et al. Herpes Simplex Virus Type 2 Seroprevalence Among Different National Populations of Middle East and North African Men. Sex Transm Dis 2018; 45(7): 482-7.
40. De Baetselier I, Menten J, Cuylaerts V, et al. Prevalence and incidence estimation of HSV-2 by two IgG ELISA methods among South African women at high risk of HIV. PloS one 2015; 10(3): e0120207.
41. Dhont N, van de Wijgert J, Luchters S, Muvunyi C, Vyankandondera J, Temmerman M. Sexual violence, HSV-2 and HIV are important predictors for infertility in Rwanda. Human reproduction (Oxford, England) 2010; 25(10): 2507-15.
42. Domercant JW, Jean Louis F, Hulland E, et al. Seroprevalence of Herpes Simplex Virus type-2 (HSV-2) among pregnant women who participated in a national HIV surveillance activity in Haiti. BMC Infect Dis 2017; 17(1): 577-.
43. Dordević H. [Serological response to herpes simplex virus type 1 and 2 infection among women of reproductive age]. Med Pregl 2006; 59(11-12): 591-7.
44. Doyle AM, Ross DA, Maganja K, et al. Long-term biological and behavioural impact of an adolescent sexual health intervention in Tanzania: follow-up survey of the community-based MEMA kwa Vijana Trial. PLoS medicine 2010; 7(6): e1000287.
45. Duflo E, Dupas P, Kremer M. Education, HIV, and Early Fertility: Experimental Evidence from Kenya. The American economic review 2015; 105(9): 2757-97.
46. Fearon E, Wiggins RD, Pettifor AE, et al. Associations between friendship characteristics and HIV and HSV-2 status amongst young South African women in HPTN-068. J Int AIDS Soc 2017; 20(4).
47. Francis SC, Mthiyane TN, Baisley K, et al. Prevalence of sexually transmitted infections among young people in South Africa: A nested survey in a health and demographic surveillance site. PLoS medicine 2018; 15(2): e1002512.
48. Gabster A, Pascale JM, Cislaghi B, et al. High Prevalence of Sexually Transmitted Infections, and High-Risk Sexual Behaviors Among Indigenous Adolescents of the Comarca Ngäbe-Buglé, Panama. Sex Transm Dis 2019; 46(12): 780-7.
49. Glynn JR, Kayuni N, Gondwe L, Price AJ, Crampin AC. Earlier menarche is associated with a higher prevalence of Herpes simplex type-2 (HSV-2) in young women in rural Malawi. eLife 2014; 3: e01604.
50. Goncalez TT, Sabino EC, Murphy EL, Chen S, Chamone DAF, McFarland W. Human immunodeficiency virus test-seeking motivation in blood donors, São Paulo, Brazil. Vox Sang 2006; 90(3): 170-6.
51. Gorfinkel IS, Aoki F, McNeil S, et al. Seroprevalence of HSV-1 and HSV-2 antibodies in Canadian women screened for enrolment in a herpes simplex virus vaccine trial. International Journal of STD and AIDS 2013; 24(5): 345-9.
52. Gray GE, Allen M, Moodie Z, et al. Safety and efficacy of the HVTN 503/Phambili study of a clade-B-based HIV-1 vaccine in South Africa: a double-blind, randomised, placebo-controlled test-of-concept phase 2b study. The Lancet Infectious diseases 2011; 11(7): 507-15.
53. Guwatudde D, Wabwire-Mangen F, Eller LA, et al. Relatively low HIV infection rates in rural Uganda, but with high potential for a rise: a cohort study in Kayunga District, Uganda. PloS one 2009; 4(1): e4145.
54. Hallfors DD, Cho H, Mbai, II, et al. Disclosure of HSV-2 serological test results in the context of an adolescent HIV prevention trial in Kenya. Sex Transm Infect 2015; 91(6): 395-400.
55. Han L, Husaiyin S, Wang X, Rouzi H, Niyazi M. Prevalence and risk factors of cervical cancer in the Nanjiang area of the Xinjiang Uyghur Autonomous Region of China: a matched case-control study. European Journal of Gynaecological Oncology 2021; 42(6): 1270-6.
56. Hazel A, Foxman B, Low BS. Herpes simplex virus type 2 among mobile pastoralists in northwestern Namibia. Annals of human biology 2015; 42(6): 543-51.
57. He N, Cao H, Yin Y, Gao M, Zhang T, Detels R. Herpes simplex virus-2 infection in male rural migrants in Shanghai, China. Int J STD AIDS 2009; 20(2): 112-4.
58. Herrera-Ortiz A, Conde-Glez CJ, Vergara-Ortega DN, García-Cisneros S, Olamendi-Portugal ML, Sánchez-Alemán MA. Avidity of antibodies against HSV-2 and risk to neonatal transmission among Mexican pregnant women. Infect Dis Obstet Gynecol 2013; 2013: 140142-.
59. Hochberg CH, Schneider JA, Dandona R, et al. Population and dyadic-based seroincidence of herpes simplex virus-2 and syphilis in southern India. Sex Transm Infect 2015; 91(5): 375-82.
60. Hokororo A, Kihunrwa A, Hoekstra P, et al. High prevalence of sexually transmitted infections in pregnant adolescent girls in Tanzania: a multi-community cross-sectional study. Sex Transm Infect 2015; 91(7): 473-8.
61. Hussan BM. Study the Prevalence of ACL, APL, CMV, HSV, Rubella and Toxoplasma Gondii in Aborted Women in Baghdad. Medical Journal of Babylon 2013; 10(2): 455-64.
62. Huai P, Li F, Li Z, et al. Seroprevalence and associated factors of HSV-2 infection among general population in Shandong Province, China. BMC Infect Dis 2019; 19(1): 382.
63. Jespers V, Crucitti T, Menten J, et al. Prevalence and correlates of bacterial vaginosis in different sub-populations of women in sub-Saharan Africa: a cross-sectional study. PloS one 2014; 9(10): e109670.
64. Jozani ZB, Badie BM, Bayanolhagh S, et al. Knowledge, attitude, and practice towards HIV/AIDS conjoint with HIV, HBV, HCV and HSV2 serosurveys among girls from dysfunctional families in Tehran, Iran. Journal of International Translational Medicine 2019; 7(1): 33-8.
65. Kane CT, Diawara S, Ndiaye HD, et al. Concentrated and linked epidemics of both HSV-2 and HIV-1/HIV-2 infections in Senegal: public health impacts of the spread of HIV. Int J STD AIDS 2009; 20(11): 793-6.
66. Karaer A, Mert I, Cavkaytar S, Batioglu S. Serological investigation of the role of selected sexually transmitted infections in the aetiology of ectopic pregnancy. The European journal of contraception & reproductive health care: the official journal of the European Society of Contraception 2013; 18(1): 68-74.
67. Kharsany ABM, McKinnon LR, Lewis L, et al. Population prevalence of sexually transmitted infections in a high HIV burden district in KwaZulu-Natal, South Africa: Implications for HIV epidemic control. International Journal of Infectious Diseases 2020; 98: 130-7.
68. Korr G, Thamm M, Czogiel I, Poethko-Mueller C, Bremer V, Jansen K. Decreasing seroprevalence of herpes simplex virus type 1 and type 2 in Germany leaves many people susceptible to genital infection: time to raise awareness and enhance control. BMC Infect Dis 2017; 17(1): 471.
69. Kucera P, Gerber S, Marques-Vidal P, Meylan PR. Seroepidemiology of herpes simplex virus type 1 and 2 in pregnant women in Switzerland: an obstetric clinic-based study. European journal of obstetrics, gynecology, and reproductive biology 2012; 160(1): 13-7.
70. Kuteesa MO, Weiss HA, Cook S, et al. Epidemiology of alcohol misuse and illicit drug use among young people aged 15-24 years in fishing communities in Uganda. International Journal of Environmental Research and Public Health 2020; 17 (7) (no pagination) (2401).
71. LeGoff J, Weiss HA, Gresenguet G, et al. Cervicovaginal HIV-1 and herpes simplex virus type 2 shedding during genital ulcer disease episodes. Aids 2007; 21(12): 1569-78.
72. Li JM, Chen YR, Li XT, Xu WC. Screening of Herpes simplex virus 2 infection among pregnant women in southern China. J Dermatol 2011; 38(2): 120-4.
73. Li Z, Yan R, Yan C, Liu P, Feng Z. Evaluation of an Automated Chemiluminescent Immunoassay in Typing Detection of IgG Antibodies Against Herpes Simplex Virus. J Clin Lab Anal 2016; 30(5): 577-80.
74. Lin H, He N, Su M, Feng J, Chen L, Gao M. Herpes simplex virus infections among rural residents in eastern China. BMC Infect Dis 2011; 11: 69.
75. Luseno WK, Hallfors DD, Cho H, et al. Use of HIV and HSV-2 biomarkers in sub-saharan adolescent prevention research: a comparison of two approaches. The journal of primary prevention 2014; 35(3): 181-91.
76. Madhivanan P, Chen YH, Krupp K, Arun A, Klausner JD, Reingold AL. Incidence of herpes simplex virus type 2 in young reproductive age women in Mysore, India. Indian J Pathol Microbiol 2011; 54(1): 96-9.
77. Marchi S, Trombetta CM, Gasparini R, Temperton N, Montomoli E. Epidemiology of herpes simplex virus type 1 and 2 in Italy: a seroprevalence study from 2000 to 2014. Journal of preventive medicine and hygiene 2017; 58(1): E27-e33.
78. Mawak JD, Dashe N, Atseye AB, Agabi YA, Zakeri H. Seroprevalence and co-infection of herpes simplex virus type 2 and human immunodeficiency virus in Nigeria. Shiraz E Medical Journal 2012; 13(1): 33-9.
79. Mehta SD, Nordgren RK, Agingu W, et al. Sexual Quality of Life and Association with HIV and Sexually Transmitted Infections Among a Cohort of Heterosexual Couples in Kenya. Journal of Sexual Medicine 2018; 15(10): 1446-55.
80. Memish ZA, Almasri M, Chentoufi AA, et al. Seroprevalence of Herpes Simplex Virus Type 1 and Type 2 and Coinfection with HIV and Syphilis: The First National Seroprevalence Survey in Saudi Arabia. Sex Transm Dis 2015; 42(9): 526-32.
81. Menezes LJ, Pokharel U, Sudenga SL, et al. Patterns of prevalent HPV and STI co-infections and associated factors among HIV-negative young Western Cape, South African women: the EVRI trial. Sex Transm Infect 2018; 94(1): 55-61.
82. Mensch BS, Grant MJ, Soler-Hampejsek E, Kelly CA, Chalasani S, Hewett PC. Does schooling protect sexual health? The association between three measures of education and STIs among adolescents in Malawi. Population studies 2020; 74(2): 241-61.
83. Mir AM, Wajid A, Reichenbach L, Khan M, Masood I. Herpes Simplex Virus-2 infection amongst urban male population in Pakistan. J Pak Med Assoc 2010; 60(11): 918-22.
84. Miskulin M, Miskulin I, Milas J, Antolovic-Pozgain A, Rudan S, Vuksic M. Prevalence and risk factors for herpes simplex virus type 2 infections in East Croatia. Collegium antropologicum 2011; 35(1): 9-14.
85. MOH Uganda, ORC Macro. Uganda HIV/AIDS sero-behavioural survey: 2004-2005: Calverton, Maryland, USA: Ministry of Health and ORC Macro., 2006.
86. Moreira-Soto A, Cabral R, Pedroso C, et al. Exhaustive TORCH Pathogen Diagnostics Corroborate Zika Virus Etiology of Congenital Malformations in Northeastern Brazil. mSphere 2018; 3(4).
87. Munawwar A, Gupta S, Sharma SK, Singh S. Seroprevalence of HSV-1 and 2 in HIV-infected males with and without GUD: Study from a tertiary care setting of India. Journal of laboratory physicians 2018; 10(3): 326-31.
88. Munro HL, Pradeep BS, Jayachandran AA, et al. Prevalence and determinants of HIV and sexually transmitted infections in a general population-based sample in Mysore district, Karnataka state, southern India. Aids 2008; 22: S117-S25.
89. Nag S, Sarkar S, Chattopadhyay D, Bhattacharya S, Biswas R, SenGupta M. Seroprevalence of Herpes Simplex Virus Infection in HIV Coinfected Individuals in Eastern India with Risk Factor Analysis. Advances in virology 2015; 2015: 537939.
90. Nakku-Joloba E, Kambugu F, Wasubire J, et al. Sero-prevalence of herpes simplex type 2 virus (HSV-2) and HIV infection in Kampala, Uganda. African health sciences 2014; 14(4): 782-9.
91. Nakubulwa S, Kaye DK, Bwanga F, Tumwesigye NM, Mirembe FM. Genital infections and risk of premature rupture of membranes in Mulago Hospital, Uganda: a case control study. BMC research notes 2015; 8: 573.
92. Nakubulwa S, Kaye DK, Bwanga F, Tumwesigye NM, Nakku-Joloba E, Mirembe FM. Incidence and risk factors for herpes simplex virus type 2 seroconversion among pregnant women in Uganda: A prospective study. Journal of infection in developing countries 2016; 10(10): 1108-15.
93. NASCOP (National AIDS/STI Control Program). 2007 Kenya AIDS Indicator Survey: Final Report. Nairobe, Kenya, September 2009.
94. Nasrallah G, Dargham S, Harfouche M, Abu-Raddad L. Seroprevalence of Herpes simplex virus types 1 and 2 in Indian and Filipino migrant populations in Qatar: a cross-sectional survey. Eastern Mediterranean health journal = La revue de sante de la Mediterranee orientale = al-Majallah al-sihhiyah li-sharq al-mutawassit 2020; 26(5): 609-15.
95. Norris AH, Kitali AJ, Worby E. Alcohol and transactional sex: how risky is the mix? Social science & medicine (1982) 2009; 69(8): 1167-76.
96. Nyiro JU, Sanders EJ, Ngetsa C, et al. Seroprevalence, predictors and estimated incidence of maternal and neonatal Herpes Simplex Virus type 2 infection in semi-urban women in Kilifi, Kenya. BMC Infect Dis 2011; 11: 155.
97. Oliver VO, Otieno G, Gvetadze R, et al. High prevalence of sexually transmitted infections among women screened for a contraceptive intravaginal ring study, Kisumu, Kenya, 2014. International Journal of STD and AIDS 2018; 29(14): 1390-9.
98. Otieno FO, Ndivo R, Oswago S, et al. Correlates of prevalent sexually transmitted infections among participants screened for an HIV incidence cohort study in Kisumu, Kenya. Int J STD AIDS 2015; 26(4): 225-37.
99. Papadogeorgakis H, Caroni C, Katsambas A, et al. Herpes simplex virus seroprevalence among children, adolescents and adults in Greece. Int J STD AIDS 2008; 19(4): 272-8.
100. Pascoe SJ, Langhaug LF, Mavhu W, et al. Poverty, food insufficiency and HIV infection and sexual behaviour among young rural Zimbabwean women. PloS one 2015; 10(1): e0115290.
101. Patzi-Churqui M, Terrazas-Aranda K, Liljeqvist JA, Lindh M, Eriksson K. Prevalence of viral sexually transmitted infections and HPV high-risk genotypes in women in rural communities in the Department of La Paz, Bolivia. BMC Infect Dis 2020; 20(1): 204.
102. Paz-Bailey G, Morales-Miranda S, Jacobson JO, et al. High rates of STD and sexual risk behaviors among Garífunas in Honduras. J Acquir Immune Defic Syndr 2009; 51 Suppl 1: S26-34.
103. Price J, Pettifor A, Selin A, et al. The association between perceived household educational support and HIV risk in young women in a rural South African community (HPTN 068): A cross sectional study. PloS one 2019; 14(1): e0210632.
104. Rahman S, Wathington D, Waterboer T, et al. Seroprevalence of Chlamydia trachomatis, herpes simplex 2, Epstein-Barr virus, hepatitis C and associated factors among a cohort of men ages 18–70 years from three countries. PloS one 2021; 16(6): e0253005.
105. Rajaram SP, Bradley JE, Ramesh BM, et al. P1-S1.14 Is HIV prevalence declining in Southern India? Evidence from two rounds of general population surveys in Bagalkot District, Karnataka. Sexually Transmitted Infections 2011; 87(Suppl 1): A105-A.
106. Rathore S, Jamwal A, Gupta V. Herpes simplex virus type 2: Seroprevalence in antenatal women. Indian journal of sexually transmitted diseases 2010; 31(1): 11.
107. Remis RS, Liu J, Loutfy M, et al. The epidemiology of sexually transmitted co-infections in HIV-positive and HIV-negative African-Caribbean women in Toronto. BMC Infect Dis 2013; 13: 550.
108. Rostamzadeh Khameneh Z, Sepehrvand N, Mohammadian M. Herpes Simplex Virus Type 2 Seroprevalence in Pregnant Women in Urmia, Northwest of Iran, during 2014-2015. Iran Biomed J 2020; 24(2): 136-9.
109. Rotermann M, Langlois KA, Severini A, Totten S. Prevalence of Chlamydia trachomatis and herpes simplex virus type 2: Results from the 2009 to 2011 Canadian Health Measures Survey. Health reports 2013; 24(4): 10-5.
110. Sasadeusz JJ, Silvers JE, Kent HE, Devenish W, Hocking J, Garland SM. Prevalence of HSV-2 antibody in a Melbourne antenatal population attending a tertiary obstetric hospital. The Australian & New Zealand journal of obstetrics & gynaecology 2008; 48(3): 266-72.
111. Schneider JA, Lakshmi V, Dandona R, Kumar GA, Sudha T, Dandona L. Population-based seroprevalence of HSV-2 and syphilis in Andhra Pradesh state of India. BMC Infect Dis 2010b; 10: 59.
112. Sgaier SK, Mony P, Jayakumar S, et al. Prevalence and correlates of Herpes Simplex Virus-2 and syphilis infections in the general population in India. Sex Transm Infect 2011; 87(2): 94-100.
113. Shahraki AD, Moghim S, Akbari P. A survey on herpes simplex type 2 antibody among pregnant women in Isfahan, Iran. Journal of Research in Medical Sciences 2010; 15(4).
114. Shen JH, Huang KY, Chao-Yu C, Chen CJ, Lin TY, Huang YC. Seroprevalence of Herpes Simplex Virus Type 1 and 2 in Taiwan and Risk Factor Analysis. PloS one 2015; 10(8): e0134178.
115. Shin HS, Park JJ, Chu C, et al. Herpes simplex virus type 2 seroprevalence in Korea: rapid increase of HSV-2 seroprevalence in the 30s in the southern part. Journal of Korean medical science 2007; 22(6): 957-62.
116. Sivapalasingam S, McClelland RS, Ravel J, et al. An effective intervention to reduce intravaginal practices among HIV-1 uninfected Kenyan women. AIDS Research and Human Retroviruses 2014; 30(11): 1046-54.
117. Suligoi B, Quaglio G, Regine V, et al. Seroprevalence of HIV, HSV-2, and Treponema pallidum in the Kosovarian population. Scandinavian journal of infectious diseases 2009; 41(8): 608-13.
118. Tobian AA, Charvat B, Ssempijja V, et al. Factors associated with the prevalence and incidence of herpes simplex virus type 2 infection among men in Rakai, Uganda. The Journal of infectious diseases 2009; 199(7): 945-9.
119. Tobian AA, Kigozi G, Redd AD, et al. Male circumcision and herpes simplex virus type 2 infection in female partners: a randomized trial in Rakai, Uganda. The Journal of infectious diseases 2012; 205(3): 486-90.
120. Todd CS, Nasir A, Mansoor GF, et al. Cross-sectional assessment of prevalence and correlates of blood-borne and sexually transmitted infections among Afghan National Army recruits. BMC Infect Dis 2012; 12: 196.
121. Topbas M, Can E, Kaklikkaya N, Yavuzyilmaz A, Ozkan G, Can G. Herpes simplex virus type-2 seroprevalence among adults aged 20-49 in Trabzon. Nobel Medicus 2012; 8(2): 85-90.
122. Vallely A, Ryan CE, Allen J, et al. High prevalence and incidence of HIV, sexually transmissible infections and penile foreskin cutting among sexual health clinic attendees in Papua New Guinea. Sex Health 2014; 11(1): 58-66.
123. Vallely AJ, MacLaren D, David M, et al. Dorsal longitudinal foreskin cut is associated with reduced risk of HIV, syphilis and genital herpes in men: a cross-sectional study in Papua New Guinea. J Int AIDS Soc 2017; 20(1): 21358.
124. Vallely LM, Toliman P, Ryan C, et al. Prevalence and risk factors of Chlamydia trachomatis, Neisseria gonorrhoeae, Trichomonas vaginalis and other sexually transmissible infections among women attending antenatal clinics in three provinces in Papua New Guinea: a cross-sectional survey. Sex Health 2016; 13(5): 420-7.
125. Vilibić-Čavlek T, Kolarić B, Bogdanić M, Tabain I, Beader N. Herpes Group Viruses: a Seroprevalence Study in Hemodialysis Patients. Acta Clin Croat 2017; 56(2): 255-61.
126. Vilibic-Cavlek T, Ljubin-Sternak S, Ban M, Kolaric B, Sviben M, Mlinaric-Galinovic G. Seroprevalence of TORCH infections in women of childbearing age in Croatia. The journal of maternal-fetal & neonatal medicine: the official journal of the European Association of Perinatal Medicine, the Federation of Asia and Oceania Perinatal Societies, the International Society of Perinatal Obstet 2011; 24(2): 280-3.
127. Wang LC, Yan F, Ruan JX, Xiao Y, Yu Y. TORCH screening used appropriately in China? ─three years results from a teaching hospital in northwest China. BMC pregnancy and childbirth 2019; 19(1): 484.
128. Warnecke JM, Pollmann M, Borchardt-Loholter V, et al. Seroprevalences of antibodies against ToRCH infectious pathogens in women of childbearing age residing in Brazil, Mexico, Germany, Poland, Turkey and China. Epidemiology and Infection 2020.
129. Winston SE, Chirchir AK, Muthoni LN, et al. Prevalence of sexually transmitted infections including HIV in street-connected adolescents in western Kenya. Sex Transm Infect 2015; 91(5): 353-9.
130. Woestenberg PJ, Tjhie JH, de Melker HE, et al. Herpes simplex virus type 1 and type 2 in the Netherlands: seroprevalence, risk factors and changes during a 12-year period. BMC Infect Dis 2016; 16: 364.
131. Yegorov S, Galiwango RM, Good SV, et al. Schistosoma mansoni infection and socio-behavioural predictors of HIV risk: a cross-sectional study in women from Uganda. BMC Infect Dis 2018; 18(1): 586.
132. Zhang JF, Zhang WY. [Relationship of cytomegalovirus, Chlamydia pneumoniae and herpes simplex virus type 2 infections with preeclampsia]. Zhonghua Yi Xue Za Zhi 2012; 92(20): 1413-5.
133. Zhang T, Yang Y, Yu F, et al. Kaposìs sarcoma associated herpesvirus infection among female sex workers and general population women in Shanghai, China: a cross-sectional study. BMC Infect Dis 2014; 14: 58.
134. Zulaika G, van Eijk AM, Mason L, et al. Factors associated with the prevalence of HIV, HSV-2, pregnancy, and reported sexual activity among adolescent girls in rural western Kenya: A cross-sectional analysis of baseline data in a cluster randomized controlled trial. PLoS Medicine 2021; 18(9): e1003756.

Abbreviations: HSV = Herpes simplex virus.

### Box S5.

List of articles on the general population from which an HSV-1 prevalence measure was extracted and included in the meta-analyses.

1. Abbas AH, KrikorMelconian A, Ad’hiah AH. No etiological role of Herpes Simplex Virus and Toxoplasma Gondii infections in systemic lupus erythematosus of Iraqi Female Patients. Biochemical and Cellular Archives 2019; 19(1): 919-21.
2. Al-Hakami AM, Paul E, Al-Abed F, et al. Prevalence of toxoplasmosis, rubella, cytomegalovirus, and herpes (TORCH) infections among women attending the antenatal care clinic, maternity hospital in Abha, Southwestern Saudi Arabia. Saudi Med J 2020; 41(7): 757-62.
3. Al-Hatemi LA, Kadhim RA, Al-Rubaye AF. Relationship of rheumatoid arthritis with microbial pathogens (TORCH Test), case-control study in Iraq. Journal of Cardiovascular Disease Research 2020; 11(4): 230-4.
4. Al-Shuwaikh AMA, Hanna DB, Ali ZQ. Seroprevalence of herpes simplex Virus type-1 IgG Antibody in Healthy Blood Donor from Baghdad, Iraq. Journal of Pure and Applied Microbiology 2019; 13(2): 1017-23.
5. Althaqafi RMM, Elrewiny M, Abdel-Moneim AS. Maternal and neonatal infections of herpes simplex virus-1 and cytomegalovirus in Saudi Arabia. J Infect Public Health 2020; 13(2): 313-4.
6. Arama V, Cercel AS, Vladareanu R, et al. Type-specific herpes simplex virus-1 and herpes simplex virus-2 seroprevalence in Romania: comparison of prevalence and risk factors in women and men. International journal of infectious diseases: IJID : official publication of the International Society for Infectious Diseases 2010; 14 Suppl 3: e25-31.
7. Arama V, Vladareanu R, Mihailescu R, et al. Seroprevalence and risk factors associated with herpes simplex virus infection among pregnant women. Journal of perinatal medicine 2008; 36(3): 206-12.
8. Ashley-Morrow R, Nollkamper J, Robinson NJ, Bishop N, Smith J. Performance of focus ELISA tests for herpes simplex virus type 1 (HSV-1) and HSV-2 antibodies among women in ten diverse geographical locations. Clinical microbiology and infection: the official publication of the European Society of Clinical Microbiology and Infectious Diseases 2004; 10(6): 530-6.
9. Ben Fredj N, Nefzi F, Aouni M, et al. Evaluation of the implication of KIR2DL2 receptor in multiple sclerosis and herpesvirus susceptibility. Journal of Neuroimmunology 2014; 271(1-2): 30-5.
10. Ben Fredj N, Nefzi F, Aouni M, et al. Identification of human herpesviruses 1 to 8 in Tunisian multiple sclerosis patients and healthy blood donors. Journal of NeuroVirology 2012; 18(1): 12-9.
11. Benharrosh J, Dauphin H, Porcheret H, Meritet JF, Boulanger MC, Maisonneuve L. [Comparison of two ELISA tests to study the seroprevalence of herpes simplex 1 et 2 infection in a maternity near Paris]. Annales de biologie clinique 2008; 66(6): 665-70.
12. Blomstrom A, Karlsson H, Wicks S, Yang S, Yolken RH, Dalman C. Maternal antibodies to infectious agents and risk for non-affective psychoses in the offspring--a matched case-control study. Schizophrenia research 2012; 140(1-3): 25-30.
13. Bogaerts J, Ahmed J, Akhter N, et al. Sexually transmitted infections among married women in Dhaka, Bangladesh: unexpected high prevalence of herpes simplex type 2 infection. Sexually transmitted infections 2001; 77(2): 114-9.
14. Brazzale AG, Russell DB, Cunningham AL, Taylor J, McBride WJ. Seroprevalence of herpes simplex virus type 1 and type 2 among the Indigenous population of Cape York, Far North Queensland, Australia. Sex Health 2010; 7(4): 453-9.
15. Centers for Disease Control and Prevention (CDC). National Center for Health Statistics (NCHS). National Health and Nutrition Examination Survey Data. Hyattsville, MD: U.S. Department of Health and Human Services, Centers for Disease Control and Prevention. 2008.
16. Centers for Disease Control and Prevention (CDC). National Center for Health Statistics (NCHS). National Health and Nutrition Examination Survey Data. Hyattsville, MD: U.S. Department of Health and Human Services, Centers for Disease Control and Prevention. 2010.
17. Centers for Disease Control and Prevention (CDC). National Center for Health Statistics (NCHS). National Health and Nutrition Examination Survey Data. Hyattsville, MD: U.S. Department of Health and Human Services, Centers for Disease Control and Prevention, 2012.
18. Centers for Disease Control and Prevention (CDC). National Center for Health Statistics (NCHS). National Health and Nutrition Examination Survey Data. Hyattsville, MD: U.S. Department of Health and Human Services, Centers for Disease Control and Prevention. 2014.
19. Centers for Disease Control and Prevention (CDC). National Center for Health Statistics (NCHS). National Health and Nutrition Examination Survey Data. Hyattsville, MD: U.S. Department of Health and Human Services, Centers for Disease Control and Prevention, 2016.
20. Chen CY, Shen JH, Huang YC. Seroepidemiology of Epstein-Barr virus and herpes simplex virus-1 in Taiwan. International Journal of Antimicrobial Agents 2013; 42(SUPPL. 2): S135.
21. Chen L, Liu J, Shi L, et al. Seasonal influence on TORCH infection and analysis of multi-positive samples with indirect immunofluorescence assay. J Clin Lab Anal 2019; 33(4): e22828.
22. Cliff JM, King EC, Lee JS, et al. Cellular Immune Function in Myalgic Encephalomyelitis/Chronic Fatigue Syndrome (ME/CFS). Front Immunol 2019; 10: 796.
23. Conde-Glez C, Lazcano-Ponce E, Rojas R, et al. Seroprevalences of varicella-zoster virus, herpes simplex virus and cytomegalovirus in a cross-sectional study in Mexico. Vaccine 2013; 31(44): 5067-74.
24. Cowan FM, French RS, Mayaud P, et al. Seroepidemiological study of herpes simplex virus types 1 and 2 in Brazil, Estonia, India, Morocco, and Sri Lanka. Sexually transmitted infections 2003; 79(4): 286-90.
25. Dordevic H. [Serological response to herpes simplex virus type 1 and 2 infection among women of reproductive age]. Med Pregl 2006; 59(11-12): 591-7.
26. Dworzanski J, Drop B, Kliszczewska E, Polz-Dacewicz M, StrycharzDudziak M. Prevalence of Epstein-Barr virus, human papillomavirus, cytomegalovirus and herpes simplex virus type 1 in patients with diabetes mellitus type 2 in south-eastern Poland. PloS one 2019; 14(9): e0222607.
27. Ghebrekidan H, Ruden U, Cox S, Wahren B, Grandien M. Prevalence of herpes simplex virus types 1 and 2, cytomegalovirus, and varicella-zoster virus infections in Eritrea. Journal of clinical virology: the official publication of the Pan American Society for Clinical Virology 1999; 12(1): 53-64.
28. Gorfinkel IS, Aoki F, McNeil S, et al. Seroprevalence of HSV-1 and HSV-2 antibodies in Canadian women screened for enrolment in a herpes simplex virus vaccine trial. Int J STD AIDS 2013; 24(5): 345-9.
29. Halawi M, Al-Hazmi A, Aljuaid A, et al. Seroprevalence of Toxoplasma gondii, Rubella, Group A Streptococcus, CMV and HSV-1 in COVID-19 Patients with Vitamin D Deficiency. Pak J Biol Sci 2021; 24(11): 1169-74.
30. Hamdani N, Daban-Huard C, Godin O, et al. Effects of Cumulative Herpesviridae and Toxoplasma gondii Infections on Cognitive Function in Healthy, Bipolar, and Schizophrenia Subjects. The Journal of clinical psychiatry 2017; 78(1): e18-e27.
31. Hemmingsson ES, Hjelmare E, Weidung B, et al. Antiviral treatment associated with reduced risk of clinical Alzheimer’s disease-A nested case-control study. Alzheimer’s and Dementia: Translational Research and Clinical Interventions 2021; 7(1): e12187.
32. Hogrefe W, Su X, Song J, Ashley R, Kong L. Detection of herpes simplex virus type 2-specific immunoglobulin G antibodies in African sera by using recombinant gG2, Western blotting, and gG2 inhibition. Journal of clinical microbiology 2002; 40(10): 3635-40.
33. Igde AF, Igde M, Yazici Z, et al. Distribution of HSV-1 IgG antibodies by two methods comparing in Turkish atopic children. The new microbiologica 2007; 30(2): 109-12.
34. Igde M, Igde FA, Yazici Z. Herpes simplex type I infection and atopy association in Turkish children with asthma and allergic rhinitis. Iranian Journal of Allergy, Asthma and Immunology 2009; 8(3): 149-54.
35. Ihsan AK, Rajaa A-J. Evaluation of immunoglobulines versus natural salivary defense in controlling recurrent herpes simplex lesions. Journal of baghdad college of dentistry 9-63 :(4)21; 2009 دادغب نانسلاا بط ةيلك ةلجم.
36. Irschick EU, Philipp S, Shahram F, et al. Investigation of bacterial and viral agents and immune status in Behcet’s disease patients from Iran. Int J Rheum Dis 2011; 14(3): 298-310.
37. Jafarzadeh A, Nemati M, Tahmasbi M, Ahmadi P, Rezayati MT, Sayadi AR. The association between infection burden in Iranian patients with acute myocardial infarction and unstable angina. Acta Med Indones 2011; 43(2): 105-11.
38. Jansen MA, van den Heuvel D, Bouthoorn SH, et al. Determinants of Ethnic Differences in Cytomegalovirus, Epstein-Barr Virus, and Herpes Simplex Virus Type 1 Seroprevalence in Childhood. The Journal of pediatrics 2016; 170: 126-34.e1-6.
39. Jary A, Burrel S, Boutolleau D, et al. Prevalence of cervical HPV infection, sexually transmitted infections and associated antimicrobial resistance in women attending cervical cancer screening in Mali. International Journal of Infectious Diseases 2021; 108: 610-6.
40. Juhl D, Mosel C, Nawroth F, et al. Detection of herpes simplex virus DNA in plasma of patients with primary but not with recurrent infection: implications for transfusion medicine? Transfusion medicine (Oxford, England) 2010; 20(1): 38-47.
41. Karachaliou M, Waterboer T, Casabonne D, et al. The natural history of human polyomaviruses and herpesviruses in early life - The rhea birth cohort in Greece. American Journal of Epidemiology 2016; 183(7): 671-9.
42. Kasubi MJ, Nilsen A, Marsden HS, Bergstrom T, Langeland N, Haarr L. Prevalence of antibodies against herpes simplex virus types 1 and 2 in children and young people in an urban region in Tanzania. Journal of clinical microbiology 2006; 44(8): 2801-7.
43. Kaur R, Gupta N, Baveja UK. Seroprevalence of HSV1 and HSV2 infections in family planning clinic attenders. The Journal of communicable diseases 2005; 37(4): 307-9.
44. Korr G, Thamm M, Czogiel I, Poethko-Mueller C, Bremer V, Jansen K. Decreasing seroprevalence of herpes simplex virus type 1 and type 2 in Germany leaves many people susceptible to genital infection: time to raise awareness and enhance control. BMC Infect Dis 2017; 17(1): 471.
45. Kramer MA, Uitenbroek DG, Ujcic-Voortman JK, et al. Ethnic differences in HSV1 and HSV2 seroprevalence in Amsterdam, the Netherlands. Euro surveillance: bulletin Europeen sur les maladies transmissibles = European communicable disease bulletin 2008; 13(24).
46. Kucera P, Gerber S, Marques-Vidal P, Meylan PR. Seroepidemiology of herpes simplex virus type 1 and 2 in pregnant women in Switzerland: an obstetric clinic-based study. European journal of obstetrics, gynecology, and reproductive biology 2012; 160(1): 13-7.
47. LeGoff J, Gresenguet G, Gody C, et al. Performance of the BioPlex 2200 multiplexing immunoassay platform for the detection of herpes simplex virus type 2 specific antibodies in African settings. Clinical and Vaccine Immunology 2011; 18(7): 1191-3.
48. LeGoff J, Saussereau E, Boulanger MC, et al. Unexpected high prevalence of herpes simplex virus (HSV) type 2 seropositivity and HSV genital shedding in pregnant women living in an East Paris suburban area. Int J STD AIDS 2007; 18(9): 593-5.
49. Leutscher P, Jensen JS, Hoffmann S, et al. Sexually transmitted infections in rural Madagascar at an early stage of the HIV epidemic: a 6-month community-based follow-up study. Sex Transm Dis 2005; 32(3): 150-5.
50. Li C, Li Y, Yang Y, et al. The Detection and Characterization of Herpes Simplex Virus Type 1 in Confirmed Measles Cases. Sci Rep 2019; 9(1): 12785.
51. Lin H, He N, Su M, Feng J, Chen L, Gao M. Herpes simplex virus infections among rural residents in eastern China. BMC Infect Dis 2011; 11: 69.
52. Marrazzo J, Rabe L, Kelly C, et al. Herpes simplex virus (HSV) infection in the VOICE (MTN 003) study: Pre-exposure prophylaxis (PrEP) for HIV with daily use of oral tenofovir, oral tenofovir-emtricitabine, or vaginal tenofovir gel. Sexually Transmitted Infections Conference: STI and AIDS World Congress 2013; 89(no pagination).
53. Matrajt L, Gantt S, Mayer BT, et al. Virus and host-specific differences in oral human herpesvirus shedding kinetics among Ugandan women and children. Sci Rep 2017; 7(1): 13105.
54. Memish ZA, Almasri M, Chentoufi AA, et al. Seroprevalence of Herpes Simplex Virus Type 1 and Type 2 and Coinfection with HIV and Syphilis: The First National Seroprevalence Survey in Saudi Arabia. Sex Transm Dis 2015; 42(9): 526-32. doi: 10.1097/OLQ.0000000000000336.
55. Mihret W, Rinke De Wit TF, Petros B, et al. Herpes simplex virus type 2 seropositivity among urban adults in Africa: Results from two cross-sectional surveys in Addis Ababa, Ethiopia. Sexually Transmitted Diseases 2002; 29(3): 175-81.
56. Nakku-Joloba E, Kambugu F, Wasubire J, et al. Sero-prevalence of herpes simplex type 2 virus (HSV-2) and HIV infection in Kampala, Uganda. African health sciences 2014; 14(4): 782-9.
57. Nasrallah GK, Dargham SR, Mohammed LI, Abu-Raddad LJ. Estimating seroprevalence of herpes simplex virus type 1 among different Middle East and North African male populations residing in Qatar. J Med Virol 2017; 17(10): 24916.
58. Neal JD, Tobian AA, Laeyendecker O, et al. Performance of the Euroline Western blot assay in the detection of herpes simplex virus type 2 antibody in Uganda, China and the USA. Int J STD AIDS 2011; 22(6): 342-4.
59. Nissen J, Trabjerg B, Pedersen MG, et al. Herpes Simplex Virus Type 1 infection is associated with suicidal behavior and first registered psychiatric diagnosis in a healthy population. Psychoneuroendocrinology 2019; 108: 150-4.
60. Papadogeorgakis H, Caroni C, Katsambas A, et al. Herpes simplex virus seroprevalence among children, adolescents and adults in Greece. Int J STD AIDS 2008; 19(4): 272-8.
61. Patnaik P, Herrero R, Morrow RA, et al. Type-specific seroprevalence of herpes simplex virus type 2 and associated risk factors in middle-aged women from 6 countries: the IARC multicentric study. Sex Transm Dis 2007; 34(12): 1019-24.
62. Perti T, Nyati M, Gray G, et al. Frequent genital HSV-2 shedding among women during labor in Soweto, South Africa. Infectious diseases in obstetrics and gynecology 2014; 2014: 258291.
63. Pourmand D, Janbakhash A. Seroepidemiology of herpes simplex virus type one in pregnant women referring to health care centers of Kermanshah (2004). Behboob 2009; 14(1): 96-100.
64. Puhakka L, Sarvikivi E, Lappalainen M, Surcel HM, Saxen H. Decrease in seroprevalence for herpesviruses among pregnant women in Finland: cross-sectional study of three time points 1992, 2002 and 2012. Infectious diseases (London, England) 2016; 48(5): 406-10.
65. Remis RS, Liu J, Loutfy M, et al. The epidemiology of sexually transmitted co-infections in HIV-positive and HIV-negative African-Caribbean women in Toronto. BMC Infect Dis 2013; 13: 550.
66. Rezaei-Chaparpordi S, Assmar M, Amirmozafari N, et al. Seroepidemiology of herpes simplex virus type 1 and 2 in northern iran. Iran J Public Health 2012; 41(8): 75-9. Epub 2012 Aug 31.
67. Salloom Df, Saleh DS, Al-Khafaji JT. Detection of Anti-Chlamydia pneumonia IgG and Anti-HSV-1 IgG antibodies in sample of Iraqi Behcet’s disease patients Journal of Biotechnology Research Center 2014; 8(2): 34-6.
68. Shen JH, Huang KY, Chao-Yu C, Chen CJ, Lin TY, Huang YC. Seroprevalence of Herpes Simplex Virus Type 1 and 2 in Taiwan and Risk Factor Analysis, 2007. PloS one 2015; 10(8): e0134178.
69. Shivaswamy K, Thappa DM, Jaisankar T, Sujatha S. High seroprevalence of HSV-1 and HSV-2 in STD clinic attendees and non-high-risk controls: a case control study at a referral hospital in south India. Indian Journal of Dermatology, Venereology & Leprology 2005; 71(1).
70. Snijders GJLJ, van Mierlo HC, Boks MP, et al. The association between antibodies to neurotropic pathogens and bipolar disorder: A study in the Dutch Bipolar (DB) Cohort and meta-analysis. Translational Psychiatry 2019; 9(1): 311.
71. Tedla Y, Shibre T, Ali O, et al. Serum antibodies to toxoplasma gondii and herpesvidae family viruses in individuals with schizophrenia and bipolar disorder: A Case - Control study. Ethiopian Medical Journal 2011; 49(3): 211-20.
72. Thomas P, Bhatia T, Gauba D, et al. Exposure to herpes simplex virus, type 1 and reduced cognitive function. Journal of Psychiatric Research 2013; 47(11): 1680-5.
73. Vilibic-Cavlek T, Kolaric B, Bogdanic M, Tabain I, Beader N. Herpes Group Viruses: a Seroprevalence Study in Hemodialysis Patients. Acta clinica Croatica 2017; 56(2): 255-61.
74. Vilibic-Cavlek T, Kolaric B, Ljubin-Sternak S, Mlinaric-Galinovic G. Herpes simplex virus infection in the Croatian population. Scandinavian journal of infectious diseases 2011; 43(11-12): 918-22.
75. Vilibic-Cavlek T, Ljubin-Sternak S, Ban M, Kolaric B, Sviben M, Mlinaric-Galinovic G. Seroprevalence of TORCH infections in women of childbearing age in Croatia. The journal of maternal-fetal & neonatal medicine: the official journal of the European Association of Perinatal Medicine, the Federation of Asia and Oceania Perinatal Societies, the International Society of Perinatal Obstet 2011; 24(2): 280-3.
76. Wang H, Yolken RH, Hoekstra PJ, Burger H, Klein HC. Antibodies to infectious agents and the positive symptom dimension of subclinical psychosis: The TRAILS study. Schizophrenia Research 2011; 129(1): 47-51.
77. Wang L-C, Ruan J-X, Xiao Y, Yan F, Yu Y. TORCH screening used appropriately in China? - Three years results from a teaching hospital in northwest China. BMC Pregnancy and Childbirth 2019; 19(1): 484.
78. Warnecke JM, Pollmann M, Borchardt-Loholter V, et al. Seroprevalences of antibodies against ToRCH infectious pathogens in women of childbearing age residing in Brazil, Mexico, Germany, Poland, Turkey, and China. Epidemiology and Infection 2020; 148: e271.
79. Woestenberg PJ, Tjhie JH, de Melker HE, et al. Herpes simplex virus type 1 and type 2 in the Netherlands: seroprevalence, risk factors and changes during a 12-year period. BMC Infect Dis 2016; 16: 364.
80. Zangana LMM. Seroprevalence of Herpes Simplex Virus type-1antibodies (IgM,IgG) in smokers in Kirkuk city-Iraq Tikrit Journal of Pure Science 2016; 21(1): 36-40.
81. Zhang Y, Traskman-Bendz L, Janelidze S, et al. Toxoplasma gondii immunoglobulin G antibodies and nonfatal suicidal self-directed violence. The Journal of clinical psychiatry 2012; 73(8): 1069-76.
82. Ziyaeyan M, Japoni A, Roostaee MH, Salehi S, Soleimanjahi H. A serological survey of Herpes Simplex Virus type 1 and 2 immunity in pregnant women at labor stage in Tehran, Iran. Pak J Biol Sci 2007; 10(1): 148-51.

Abbreviations: HSV = Herpes simplex virus.

**Table S3.** HSV-2 prevalence measures among general populations included in the WHO HSV estimations, by region, sex, and age group.

| WHO Region | Sex | Number of data points that contributed to the estimation by age group (in years) |  |  |  |  |  |  | Countries with data |
| --- | --- | --- | --- | --- | --- | --- | --- | --- | --- |
|  |  | 15-19 | 20-24 | 25-29 | 30-34 | 35-39 | 40-44 | 45-49 |  |
| African | Females | 45 | 33 | 27 | 19 | 9 | 8 | 4 | Benin, Côte d'Ivoire, Ethiopia, Kenya, Malawi, Namibia, Nigeria, Rwanda, Senegal, South Africa, Tanzania, Uganda, Zambia, Zimbabwe |
|  | Males | 17 | 22 | 10 | 11 | 5 | 10 | 4 | Benin, Kenya, Malawi, Namibia, Nigeria, Rwanda, South Africa, Tanzania, Uganda, Zambia |
| Americas | Females | 7 | 8 | 11 | 11 | 7 | 8 | 7 | Bolivia, Brazil, Canada, Haiti, Honduras, Mexico, Panama, USA |
|  | Males | 6 | 6 | 6 | 6 | 6 | 11 | 6 | Brazil, Canada, Honduras, Mexico, Panama, USA |
| Eastern Mediterranean | Females | 1 | 1 | 4 | 6 | 1 | - | - | Iran, Iraq, Pakistan, Saudi Arabia, Sudan |
|  | Males | 4 | 2 | 10 | 5 | 8 | 5 | 5 | Afghanistan, Egypt, Iraq, Jordan, Lebanon, Pakistan, Palestine, Qatar, Saudi Arabia, Syria |
| European | Females | 6 | 11 | 15 | 16 | 10 | 7 | 6 | Croatia, Finland, France, Germany, Greece, Italy, Netherlands, Norway, Poland, Romania, Russia, Serbia, Sweden, Switzerland, Turkey |
|  | Males | 3 | 9 | 7 | 10 | 8 | 6 | 5 | Croatia, Germany, Greece, Italy, Netherlands, Norway, Romania, Russia, Serbia, Sweden, Turkey |
| South-East Asia | Females | 4 | 6 | 8 | 4 | 4 | 3 | 1 | India |
|  | Males | 1 | 3 | 4 | 6 | 3 | 4 | 2 | India |
| Western Pacific | Females | 2 | 7 | 12 | 12 | 6 | 5 | 7 | Australia, China, Korea, Papua New Guinea, Vietnam |
|  | Males | 2 | 5 | 6 | 8 | 4 | 5 | 4 | Australia, China, Korea, Papua New Guinea, Philippines |

Abbreviations: HSV= Herpes simplex virus, USA = United States of America, WHO = World Health Organization. Regions are per World Health Organization definitions.

**Table S4.** HSV-1 prevalence measures among general populations included in the WHO HSV estimations, by region, sex, and age group.

| WHO Region | Sex | Number of data points that contributed to the estimation by age group (in years) |  |  |  |  |  |  |  |  |  | Countries with data |
| --- | --- | --- | --- | --- | --- | --- | --- | --- | --- | --- | --- | --- |
|  |  | 0-4 | 5-9 | 10-14 | 15-19 | 20-24 | 25-29 | 30-34 | 35-39 | 40-44 | 45-49 |  |
| **African** | Females, males, and mixed sexes | 3 | 2 | 3 | 4 | - | 6 | 7 | 2 | 2 | - | Central African Republic, Mali, South Africa, Uganda, Zimbabwe |
| **Americas** | Females | - | 2<sup>a</sup> | 10<sup>a</sup> | 6 | 6 | 8 | 9 | 6 | 5 | 5 | Brazil, Canada, Mexico, USA |
|  | Males |  |  |  | 6 | 5 | 6 | 6 | 6 | 5 | 5 | Mexico, USA |
| **Eastern Mediterranean** | Females, males, and mixed sexes | - | - | - | 3 | 4 | 8 | 7 | 17 | 7 | 5 | Egypt, Iran, Iraq, Jordan, Lebanon, Morocco, Pakistan, Palestine, Qatar, Sudan, Syria, Tunisia, Yemen |
| **European** | Females | 7<sup>a</sup> | 6<sup>a</sup> | 5<sup>a</sup> | 4 | 6 | 10 | 10 | 5 | 5 | 2 | Croatia, Finland, France, Germany, Greece, Netherland, Poland, Romania, Serbia, Sweden, Switzerland, Turkey |
|  | Males |  |  |  | 2 | 3 | 4 | 3 | 3 | 3 | 2 | Germany, Greece, Netherland, Romania, Sweden |
| **South-East Asia** | Females, males, and mixed sexes | - | 1 | - | 4 | 2 | 3 | 8 | 2 | 4 | 1 | India, Indonesia |
| **Western Pacific** | Females, males, and mixed sexes | 2 | 12 | 5 | 2 | 5 | 4 | 15 | 3 | 6 | 5 | Australia, China, Philippines |

Abbreviations: HSV = Herpes simplex virus, USA = United States of America, WHO = World Health Organization.
<sup>a</sup>These measures included females, males, and mixed sexes for given age bands.
Regions are per World Health Organization definitions.

**Table S5.** Comparison of HSV estimates over the WHO HSV estimation rounds [15, 17–19].

| 2012 global estimates | 2016 global estimates | 2020 global estimates |
| --- | --- | --- |
| • 37 articles were used as modelling input for HSV-1<br>• 111 articles were used as modelling input for HSV-2 | • 44 articles were used as modelling input for HSV-1<br>• 88 articles were used as modelling input for HSV-2 | • 82 articles were used as modelling input for HSV-1<br>• 134 articles were used as modelling input for HSV-2 |
| **HSV-1 incidence at any site (oral and genital):**<br>• 118.0 million new HSV-1 infections among individuals aged 0-49 years<br>• HSV-1 incidence rate of 2.0% | **HSV-1 incidence at any site (oral and genital):**<br>• 120.4 million new HSV-1 infections among individuals aged 0-49 years<br>• HSV-1 incidence rate of 2.1% | **HSV-1 incidence at any site (oral and genital):**<br>• 122.2 million new HSV-1 infections among individuals aged 0-49 years<br>• HSV-1 incidence rate of 2.1% |
| **HSV-1 prevalence at any site (oral and genital):**<br>• 3,709.0 million HSV-1 prevalent infections among individuals aged 0-49 years<br>• HSV-1 prevalence of 67.0% | **HSV-1 prevalence at any site (oral and genital):**<br>• 3,752.0 million HSV-1 prevalent infections among individuals aged 0-49 years<br>• HSV-1 prevalence of 66.6% | **HSV-1 prevalence at any site (oral and genital):**<br>• 3,779.1 million HSV-1 prevalent infections among individuals aged 0-49 years<br>• HSV-1 prevalence of 64.2% |
| **Oral HSV-1 prevalence:**<br>No estimates were produced | **Oral HSV-1 prevalence:**<br>• 3,583.5 million oral HSV-1 prevalent infections among individuals aged 0-49 years<br>• Oral HSV-1 prevalence of 63.6% | **Oral HSV-1 prevalence:**<br>• 3,448.9 million oral HSV-1 prevalent infections among individuals aged 0-49 years<br>• Oral HSV-1 prevalence of 58.6% |
| **Genital HSV-1 prevalence:**<br>• 140.0 million genital HSV-1 prevalent infections among individuals aged 15-49 years<br>• Genital HSV-1 prevalence of 4.0% | **Genital HSV-1 prevalence:**<br>• 192.0 million genital HSV-1 prevalent infections among individuals aged 15-49 years<br>• Genital HSV-1 prevalence of 5.2% | **Genital HSV-1 prevalence:**<br>• 376.2 million genital HSV-1 prevalent infections among individuals aged 15-49 years<br>• Genital HSV-1 prevalence of 10.2% |
| **HSV-2 Incidence:**<br>• 19.2 million new HSV-2 infections among individuals aged 15-49 years<br>• HSV-2 incidence rate of 0.5% | **HSV-2 Incidence:**<br>• 23.9 million new HSV-2 infections among individuals aged 15-49 years<br>• HSV-2 incidence rate of 0.6% | **HSV-2 Incidence:**<br>• 25.6 million new HSV-2 infections among individuals aged 15-49 years<br>• HSV-2 incidence rate of 0.7% |
| **HSV-2 prevalence:**<br>• 417.3 million HSV-2 prevalent infections among individuals aged 15-49 years<br>• HSV-2 prevalence of 11.3% | **HSV-2 prevalence:**<br>• 491.5 million HSV-2 prevalent infections among individuals aged 15-49 years<br>• HSV-2 prevalence of 13.2% | **HSV-2 prevalence:**<br>• 519.5 million HSV-2 prevalent infections among individuals aged 15-49 years<br>• HSV-2 prevalence of 13.3% |
| **GUD caused by HSV-1:**<br>No estimates were produced | **GUD caused by HSV-1:**<br>• 9.2 million individuals living with GUD due to HSV-1 infection among individuals aged 15-49 years<br>• Prevalence of GUD caused by HSV-1 of 0.2% | **GUD caused by HSV-1:**<br>• 16.7 million individuals living with GUD due to HSV-1 infection among individuals aged 15-49 years<br>• Prevalence of GUD caused by HSV-1 of 0.5% |
| **GUD caused by HSV-2:**<br>No estimates were produced | **GUD caused by HSV-2:**<br>• 177.7 million individuals living with GUD due to HSV-2 infection among individuals aged 15-49 years<br>• Prevalence of GUD caused by HSV-2 of 4.8% | **GUD caused by HSV-2:**<br>• 187.9 million individuals living with GUD due to HSV-2 infection among individuals aged 15-49 years<br>• Prevalence of GUD caused by HSV-2 of 4.8% |

Abbreviations: GUD = Genital ulcer disease, HSV = Herpes simplex virus, WHO = World Health Organization.

**Figure S1.**
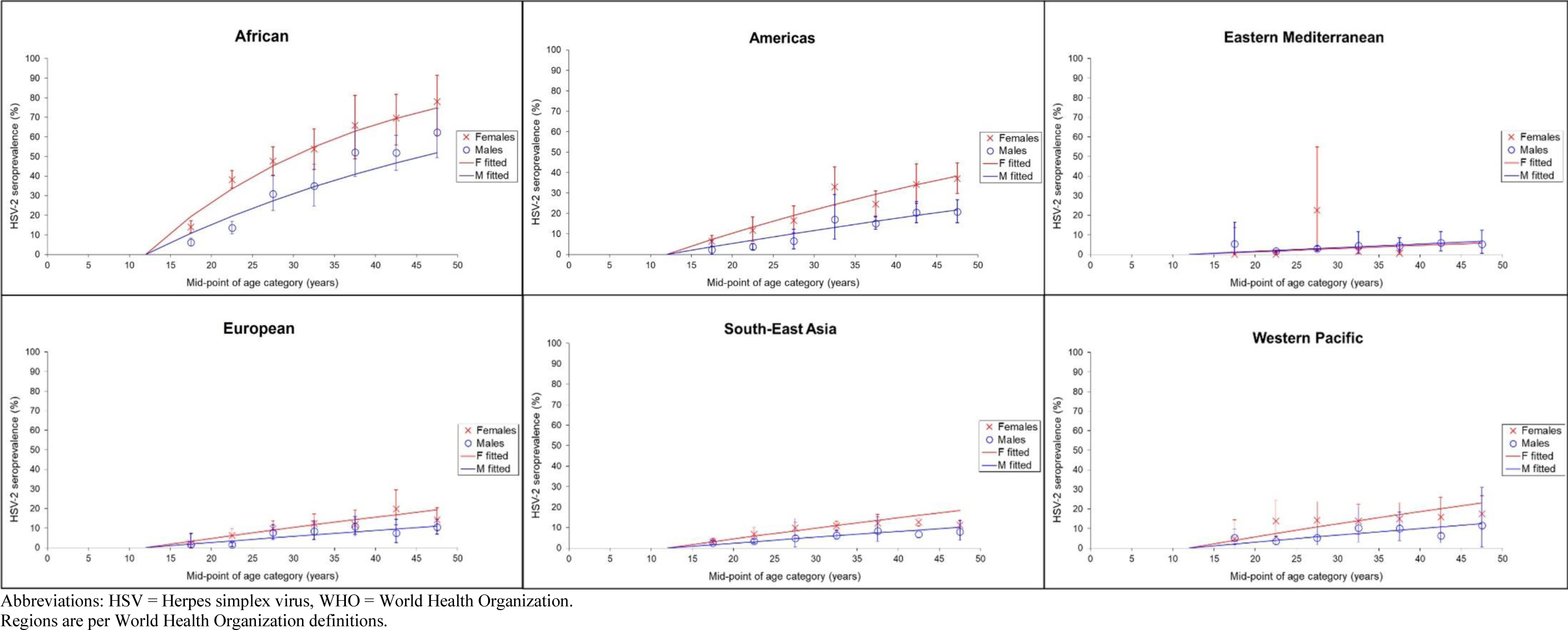
Model fits for HSV-2 prevalence for each WHO region.

**Figure S2.**
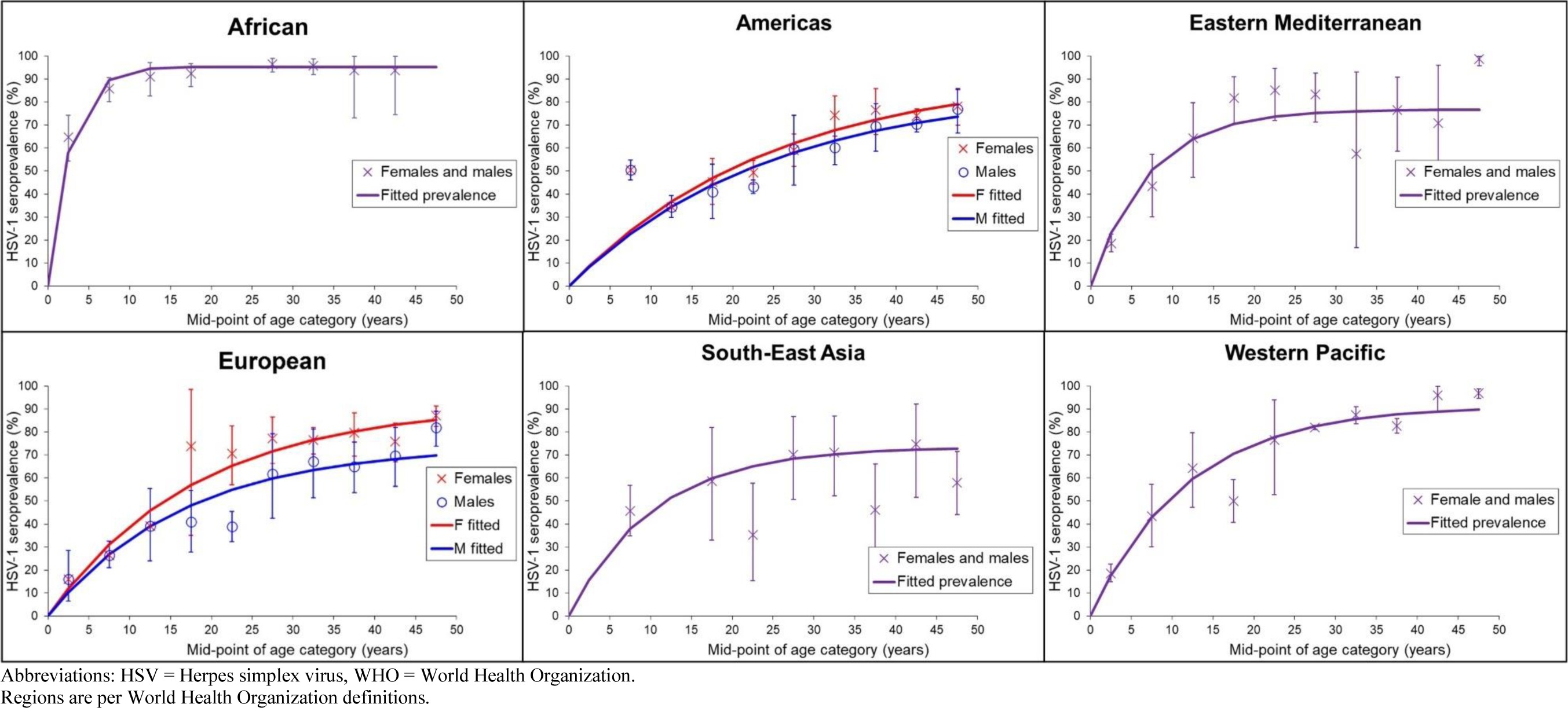
Model fits for HSV-1 prevalence (any site; oral and genital) for each WHO region.

**Table S6.** Global and regional estimates of the number of people aged 50-99 years with prevalent HSV-1 or HSV-2 infection in 2020, by sex.

| WHO Region | Number of people with prevalent infection in millions by sex |  |  |  |  |  |
| --- | --- | --- | --- | --- | --- | --- |
|  | HSV-1 (any site, oral or genital) |  |  | HSV-2 |  |  |
|  | Female | Male | Both | Female | Male | Both |
| **AFR** | 58.2 | 51.3 | **109.4** | 45.7 | 28.0 | **73.7** |
| **AMR** | 119.6 | 97.7 | **217.3** | 58.0 | 28.9 | **86.9** |
| **EMR** | 40.4 | 41.2 | **81.5** | 3.1 | 3.6 | **6.7** |
| **EUR** | 158.8 | 104.9 | **263.6** | 36.0 | 16.5 | **52.5** |
| **SEAR** | 150.1 | 145.7 | **295.7** | 37.7 | 20.3 | **58.1** |
| **WPR** | 286.0 | 270.0 | **555.9** | 73.4 | 37.8 | **111.2** |
| **Global** | **813.0** | **710.6** | **1,523.6** | **254.0** | **135.2** | **389.1** |

Abbreviations: AFR = African Region, AMR = Region of the Americas, EMR = Eastern Mediterranean Region, EUR = European Region, HSV = Herpes simplex virus, SEAR = South-East Asia region, WHO = World Health Organization, WPR = Western Pacific Region.
Numbers are the year 2020 estimated number of people living with HSV-1 and HSV-2 infections. Numbers do not always sum exactly to the totals due to rounding. Regions are per World Health Organization definitions.

**Table S7.**
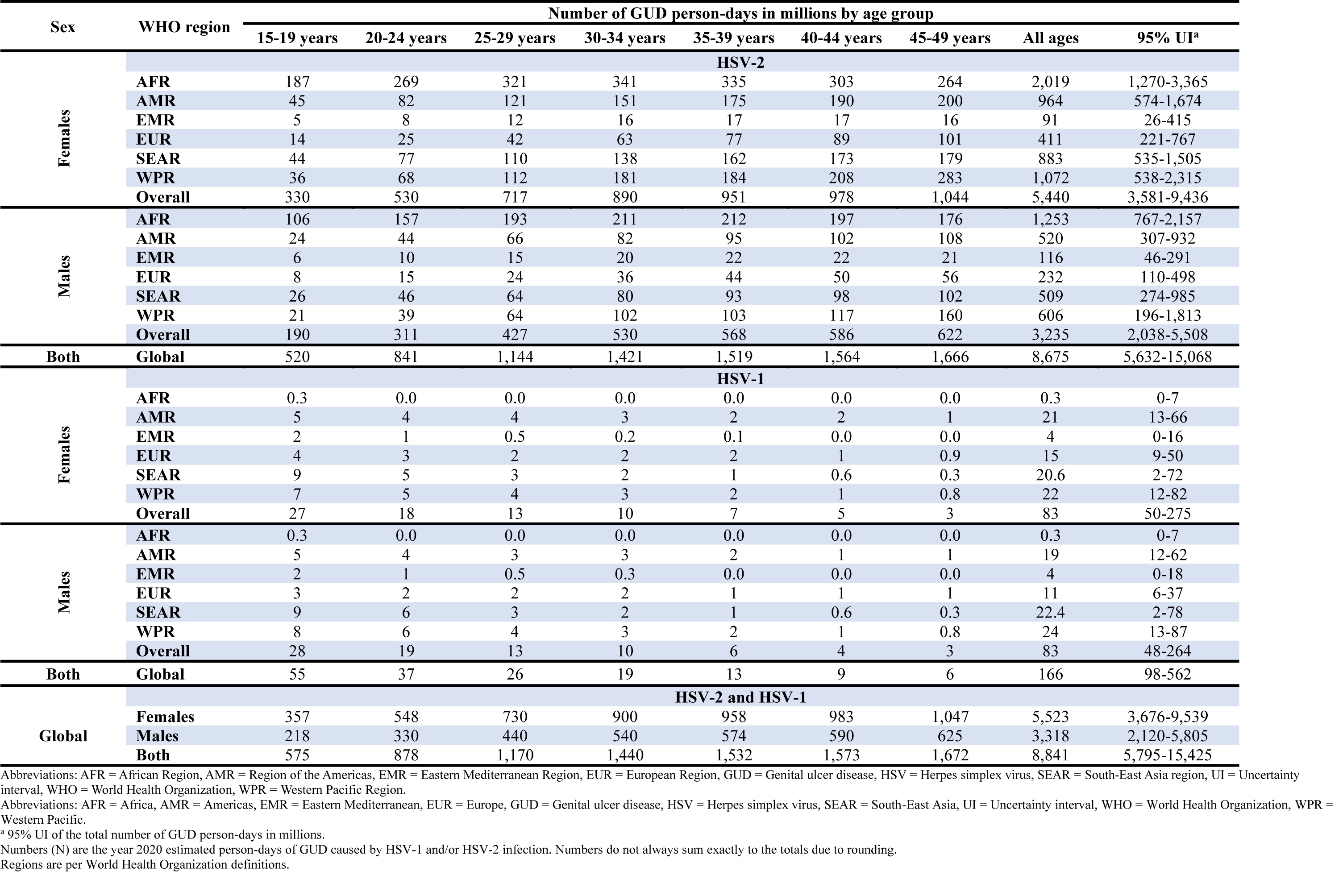
Global and regional estimates of GUD person-days due to HSV-2 or HSV-1 among the population aged 15-49 years in 2020, by age and sex.

**Table S8.**
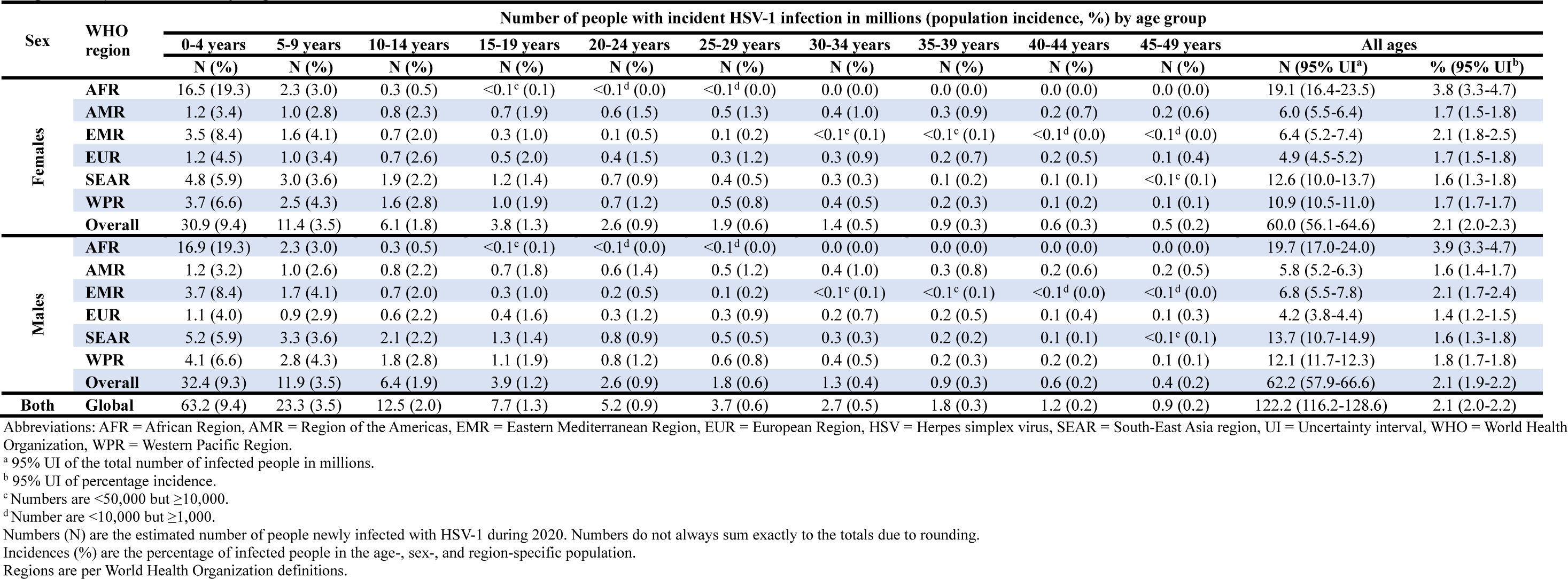
Global and regional estimates of the number and percentage of the population aged 0-49 years with incident HSV-1 infection (any site, oral or genital) in 2020, by age and sex.

**Table S9.** Global and regional estimates of the number and percentage of the population aged 0-49 years with prevalent HSV-1 infection (any site, oral or genital) in 2020, by age and sex.

| Sex | WHO region | Number of people with prevalent HSV-1 infection in millions (population prevalence, %) by age group |  |  |  |  |  |  |  |  |  |  |  |
| --- | --- | --- | --- | --- | --- | --- | --- | --- | --- | --- | --- | --- | --- |
|  |  | 0-4 years | 5-9 years | 10-14 years | 15-19 years | 20-24 years | 25-29 years | 30-34 years | 35-39 years | 40-44 years | 45-49 years | All ages |  |
|  |  | N (%) | N (%) | N (%) | N (%) | N (%) | N (%) | N (%) | N (%) | N (%) | N (%) | N (95% UI<sup>a</sup>) | % (95% UI<sup>b</sup>) |
| Females | AFR | 37.2 (57.9) | 67.1 (89.5) | 63.9 (94.4) | 55.8 (95.1) | 48.1 (95.3) | 41.5 (95.3) | 35.9 (95.3) | 30.6 (95.3) | 24.9 (95.3) | 20.1 (95.3) | 425.2 (381.9-446.7) | 85.1 (76.4-89.4) |
|  | AMR | 2.5 (8.8) | 8.2 (24.1) | 13.0 (36.6) | 17.2 (46.8) | 21.1 (55.2) | 24.2 (62.1) | 25.6 (67.7) | 26.3 (72.3) | 25.7 (76.1) | 25.0 (79.2) | 188.9 (165.2-210.9) | 51.7 (45.3-57.8) |
|  | EMR | 7.5 (23.1) | 18.8 (50.5) | 22.0 (63.9) | 22.3 (70.4) | 22.2 (73.6) | 22.1 (75.2) | 21.4 (75.9) | 19.0 (76.3) | 15.9 (76.5) | 13.0 (76.6) | 184.0 (137.9-207.1) | 61.7 (46.2-69.4) |
|  | EUR | 2.5 (11.8) | 8.2 (31.0) | 12.0 (45.7) | 14.1 (56.8) | 16.7 (65.3) | 20.9 (71.7) | 25.4 (76.6) | 26.5 (80.3) | 26.9 (83.1) | 27.1 (85.2) | 180.3 (153.4-203.9) | 61.4 (52.2-69.4) |
|  | SEAR | 10.0 (15.7) | 29.5 (37.8) | 43.2 (51.5) | 51.1 (59.9) | 54.6 (65.1) | 55.0 (68.3) | 54.5 (70.3) | 52.7 (71.6) | 47.9 (72.3) | 43.5 (72.8) | 442.0 (288.6-520.7) | 56.7 (37.0-66.8) |
|  | WPR | 7.7 (17.4) | 23.3 (42.8) | 32.2 (59.6) | 37.8 (70.5) | 44.5 (77.7) | 52.8 (82.5) | 67.9 (85.6) | 57.7 (87.6) | 55.9 (88.9) | 67.3 (89.8) | 447.0 (404.2-479.9) | 71.0 (64.2-76.2) |
|  | Overall | 67.4 (20.6) | 155.0 (48.4) | 186.3 (60.4) | 198.3 (67.4) | 207.2 (72.0) | 216.5 (75.4) | 230.7 (78.3) | 212.8 (79.8) | 197.2 (81.3) | 196.0 (82.9) | 1,867.4 (1,695.3-1,969.7) | 65.2 (59.2-68.7) |
| Males | AFR | 38.3 (57.9) | 68.8 (89.5) | 65.3 (94.4) | 56.8 (95.1) | 48.6 (95.3) | 41.6 (95.3) | 35.7 (95.3) | 30.1 (95.3) | 24.4 (95.3) | 19.5 (95.3) | 429.1 (388.5-450.8) | 84.9 (76.9-89.2) |
|  | AMR | 2.5 (8.4) | 8.0 (22.7) | 12.7 (34.4) | 16.7 (44.0) | 20.2 (51.7) | 23.1 (58.1) | 24.0 (63.2) | 24.1 (67.5) | 23.1 (70.9) | 22.2 (73.7) | 176.7 (150.9-204.5) | 47.8 (40.8-55.4) |
|  | EMR | 7.9 (23.1) | 19.9 (50.5) | 23.2 (63.9) | 23.6 (70.4) | 23.6 (73.6) | 24.1 (75.2) | 23.6 (75.9) | 20.9 (76.3) | 17.6 (76.5) | 14.3 (76.6) | 198.8 (145.8-222.6) | 61.9 (45.4-69.4) |
|  | EUR | 2.3 (10.3) | 7.5 (26.8) | 10.8 (39.1) | 12.6 (48.1) | 14.8 (54.8) | 18.2 (59.8) | 21.5 (63.5) | 22.0 (66.2) | 21.8 (68.2) | 21.7 (69.7) | 153.2 (123.7-175.4) | 50.7 (40.9-58.0) |
|  | SEAR | 10.9 (15.7) | 31.9 (37.8) | 47.0 (51.5) | 56.1 (59.9) | 59.8 (65.1) | 59.5 (68.3) | 57.7 (70.3) | 55.3 (71.6) | 49.6 (72.3) | 44.6 (72.8) | 472.5 (303.0-560.2) | 56.5 (36.3-67.0) |
|  | WPR | 8.5 (17.4) | 26.1 (42.8) | 36.6 (59.6) | 42.6 (70.5) | 49.5 (77.7) | 57.6 (82.5) | 72.0 (85.6) | 60.4 (87.6) | 58.3 (88.9) | 69.8 (89.8) | 481.4 (437.8-517.0) | 70.5 (64.1-75.7) |
|  | Overall | 70.3 (20.2) | 162.3 (47.5) | 195.7 (59.3) | 208.4 (66.1) | 216.4 (70.5) | 224.2 (73.6) | 234.6 (76.2) | 212.8 (77.4) | 194.8 (78.6) | 192.2 (80.1) | 1,911.7 (1,722.8-2,023.9) | 63.4 (57.1-67.1) |
| Both | Global | 137.7 (20.4) | 317.2 (47.9) | 382.1 (59.8) | 406.7 (66.7) | 423.6 (71.2) | 440.7 (74.5) | 465.3 (77.2) | 425.6 (78.6) | 392.0 (79.9) | 388.2 (81.5) | 3,779.1 (3,510.3-3,921.6) | 64.2 (59.7-66.7) |

Abbreviations: AFR = African Region, AMR = Region of the Americas, EMR = Eastern Mediterranean Region, EUR = European Region, HSV = Herpes simplex virus, SEAR = South-East Asia region, UI = Uncertainty interval, WHO = World Health Organization, WPR = Western Pacific Region.
<sup>a</sup> 95% UI of the total number of infected people in millions.
<sup>b</sup> 95% UI of percentage prevalence.
Numbers (N) are the year 2020 estimated number of people living with HSV-1 infection. Numbers do not always sum exactly to the totals due to rounding.
Prevalences (%) are the percentage of infected people in the age-, sex-, and region-specific population.
Regions are per World Health Organization definitions.

**Table S10.** Global and regional estimates of the number and percentage of the population aged 0-49 years with prevalent oral HSV-1 infection in 2020, by age and sex.

| Number of people with prevalent oral HSV-1 infection in millions (population prevalence, %) by age group |  |  |  |  |  |  |  |  |  |  |  |  |  |
| --- | --- | --- | --- | --- | --- | --- | --- | --- | --- | --- | --- | --- | --- |
| Sex | WHO region | 0-4 years | 5-9 years | 10-14 years | 15-19 years | 20-24 years | 25-29 years | 30-34 years | 35-39 years | 40-44 years | 45-49 years | All ages |  |
|  |  | N (%) | N (%) | N (%) | N (%) | N (%) | N (%) | N (%) | N (%) | N (%) | N (%) | N (95% UI<sup>a</sup>) | % (95% UI<sup>b</sup>) |
| Females | AFR | 37.2 (57.9) | 67.1 (89.5) | 63.9 (94.4) | 55.8 (95.0) | 48.0 (95.0) | 41.4 (95.1) | 35.9 (95.1) | 30.5 (95.1) | 24.9 (95.1) | 20.1 (95.1) | 424.7 (372.9-446.7) | 85.0 (74.6-89.4) |
|  | AMR | 2.5 (8.8) | 8.2 (24.1) | 13.0 (36.6) | 16.3 (43.7) | 18.0 (46.8) | 19.4 (49.3) | 19.5 (51.3) | 19.3 (53.0) | 18.4 (54.4) | 17.6 (55.5) | 152.2 (129.8-174.7) | 41.7 (35.6-47.9) |
|  | EMR | 7.5 (23.1) | 18.8 (50.5) | 22.0 (63.9) | 21.8 (68.7) | 21.1 (69.9) | 20.7 (70.5) | 19.9 (70.7) | 17.7 (70.9) | 14.7 (70.9) | 12.1 (71.0) | 176.3 (118.5-206.7) | 59.1 (39.7-69.3) |
|  | EUR | 2.5 (11.8) | 8.2 (31.0) | 12.0 (45.7) | 13.4 (53.5) | 14.6 (56.6) | 17.3 (58.9) | 20.2 (60.7) | 20.6 (62.1) | 20.4 (63.1) | 20.3 (63.9) | 149.5 (121.4-178.2) | 50.9 (41.3-60.6) |
|  | SEAR | 10.0 (15.7) | 29.5 (37.8) | 43.2 (51.5) | 49.5 (57.6) | 50.0 (59.5) | 49.0 (60.6) | 47.6 (61.4) | 45.6 (61.8) | 41.2 (62.1) | 37.2 (62.2) | 402.8 (231.3-516.6) | 51.7 (29.7-66.3) |
|  | WPR | 7.7 (17.4) | 23.3 (42.8) | 32.2 (59.6) | 36.5 (67.4) | 40.3 (70.0) | 46.1 (71.8) | 57.9 (72.9) | 48.5 (73.6) | 46.7 (74.1) | 55.8 (74.4) | 395.0 (340.5-443.3) | 62.7 (54.1-70.4) |
|  | Overall | 67.4 (20.6) | 155.0 (48.4) | 186.3 (60.4) | 193.3 (65.7) | 192.1 (66.8) | 193.8 (67.5) | 201.0 (68.2) | 182.2 (68.3) | 166.2 (68.5) | 163.1 (18.9) | 1,700.4 (1,501.7-1,841.0) | 59.3 (52.4-64.2) |
| Males | AFR | 38.3 (57.9) | 68.8 (89.5) | 65.3 (94.4) | 56.7 (95.0) | 48.5 (95.0) | 41.5 (95.1) | 35.6 (95.1) | 30.0 (95.1) | 24.4 (95.1) | 19.5 (95.1) | 428.6 (380.5-450.8) | 84.8 (75.3-89.2) |
|  | AMR | 2.5 (8.4) | 8.0 (22.7) | 12.7 (34.4) | 15.8 (41.1) | 17.3 (43.9) | 18.5 (46.2) | 18.3 (48.1) | 17.8 (49.6) | 16.6 (50.9) | 15.7 (51.9) | 143.3 (118.8-172.3) | 38.8 (32.2-46.6) |
|  | EMR | 7.9 (23.1) | 19.9 (50.5) | 23.2 (63.9) | 23.1 (68.7) | 22.5 (69.9) | 22.7 (70.5) | 22.0 (70.7) | 19.4 (70.9) | 16.3 (70.9) | 13.3 (71.0) | 190.3 (124.3-222.1) | 59.3 (38.7-69.2) |
|  | EUR | 2.3 (10.3) | 7.5 (26.8) | 10.8 (39.1) | 12.0 (45.4) | 13.0 (47.9) | 15.2 (49.7) | 17.4 (51.0) | 17.4 (52.0) | 16.9 (52.8) | 16.6 (53.3) | 129.0 (97.9-157.3) | 42.7 (32.4-52.1) |
|  | SEAR | 10.9 (15.7) | 31.9 (37.8) | 47.0 (51.5) | 54.4 (57.6) | 54.8 (59.5) | 53.0 (60.6) | 50.5 (61.4) | 47.8 (61.8) | 42.6 (62.1) | 38.2 (62.2) | 431.0 (242.9-556.4) | 51.6 (29.1-66.6) |
|  | WPR | 8.5 (17.4) | 26.1 (42.8) | 36.6 (59.6) | 41.1 (67.4) | 44.8 (70.0) | 50.3 (71.8) | 61.4 (72.9) | 50.8 (73.6) | 48.7 (74.1) | 57.9 (74.4) | 426.2 (371.0-477.7) | 62.4 (54.3-70.0) |
|  | Overall | 70.3 (20.2) | 162.3 (47.5) | 195.7 (59.3) | 203.2 (64.4) | 200.9 (65.5) | 201.1 (66.1) | 205.2 (66.7) | 183.2 (66.7) | 165.4 (66.8) | 161.1 (67.2) | 1,748.5 (1,531.6-1,905.2) | 58.0 (50.8-63.2) |
| Both | Global | 137.7 (20.4) | 317.2 (47.9) | 382.1 (59.8) | 396.5 (65.0) | 393.0 (66.1) | 394.9 (66.8) | 406.3 (67.4) | 365.4 (67.5) | 331.7 (67.6) | 324.2 (68.1) | 3,448.9 (3,144.9-3,655.2) | 58.6 (53.5-62.1) |

Abbreviations: AFR = African Region, AMR = Region of the Americas, EMR = Eastern Mediterranean Region, EUR = European Region, HSV = Herpes simplex virus, SEAR = South-East Asia region, UI = Uncertainty interval, WHO = World Health Organization, WPR = Western Pacific Region.
<sup>a</sup> 95% UI of the total number of infected people in millions.
<sup>b</sup> 95% UI of percentage prevalence.
Numbers (N) are the year 2020 estimated number of people living with oral HSV-1 infection. Numbers do not always sum exactly to the totals due to rounding.
Prevalences (%) are the percentage of infected people in the age-, sex-, and region-specific population.
Regions are per World Health Organization definitions.

